# Extracting Symptoms of Psychotic Disorders from Clinical Notes using Natural Language Processing

**DOI:** 10.64898/2026.09.16.26363090

**Authors:** Siobhan K. Lock, Joanne Boisson, Lowri M. Evans, Yuefeng Shi, Djenifer B. Kappel, James T.R. Walters, Michael C. O’Donovan, Sophie E. Legge, José Camacho-Collados, Antonio F. Pardiñas

## Abstract

Large Language Models (LLMs) are proposed as tools for high-throughput, deep phenotyping of psychiatric disorders. Applied to electronic health records, LLMs could in principle extract patient symptoms, outcome trajectories, risk factors, and treatment history at scale and these, when combined with increasingly available biological data, such as genomics or neuroimaging, could provide powerful resources for health research. Although proof-of-principle LLM-based symptom extractions have been carried out for some medical conditions, the heterogeneous nature of psychiatric disorders, including schizophrenia, requires extensive domain-specific evaluations. Here, we evaluate 14 general-purpose LLMs for extracting eight symptoms of schizophrenia from clinical summaries in a severe mental illness cohort (N = 704). Performance across symptoms was poor to moderate (macro F1 = 0.500 – 0.647), with positive symptoms more accurately extracted than negative symptoms. Few-shot prompting, a common strategy for improving LLM task performance, did not significantly improve these results. Nevertheless, LLM-predicted and gold-standard positive symptoms demonstrated comparable associations with clinical variables in regression analyses. Individual-level extraction errors attenuated group-level associations but did so unevenly across symptom domains. This indicates that general-purpose LLMs may not be able to extract psychosis-related phenotypes from clinical summaries with the quality required for clinical use but might nevertheless be useful for exploratory research on large cohorts. However, closing the performance gap between positive and negative symptom extractions seems essential groundwork to prevent a systematic bias in any LLM-derived characterisations of psychotic symptoms.

## Introduction

Schizophrenia is characterised by experiences of hallucinations, delusions, disorganised thoughts and behaviours, negative symptoms, and is additionally associated with abnormalities of movement and with cognitive impairment (Kahn et al. 2015). There is high inter-individual variability in the types, severity, and associated impairment of symptoms experienced (Arango and Carpenter 2010). Longitudinal assessment of symptoms is necessary to monitor both a patient’s mental health and their response to therapy. Standardised scales and questionnaires are the primary method for collecting these data in research contexts, however, their routine use in clinical settings is not common, chiefly due to time constraints (Nasrallah 2009; Aboraya et al. 2018). Yet, a trade-off remains in research between collecting detailed information and keeping data collection short enough to maximise recruitment and minimise drop-out (Edwards et al. 2023). This trade-off imposes limitations on the ability of researchers to explore symptoms longitudinally, and at scale.

The ongoing global transition from analogue to digital medical records has created a real-world database of historic interactions between patients and healthcare services (Knevel and Liao 2023). Electronic Health Records (EHRs) may contain both structured and unstructured data. Psychiatric secondary mental health care notes contain unstructured ‘free text’ records that usually holds information regarding diagnosis, symptoms, functional outcomes, medication, and case history that could in principle support analyses or enhance existing datasets (Smoller 2018). These descriptions are not equivalent to systematic questionnaire-derived symptom ratings. Instead, they contain qualitative descriptions of the patients’ experiences and care, through the lens of their clinical team and the context they operate within (Aaslestad and Skuggevik 2018).

There are, on average, eight symptoms described per psychiatric note (Forbush et al. 2013), and these are buried within the unstructured text of EHRs. These textual data often contain repetitions, temporal inconsistencies, biases, missingness, and other inaccuracies which can make symptom extraction a challenge (Grzenda and Widge 2024; Aaslestad et al. 2026). Natural Language Processing (NLP) has been used to try to address the challenges of translating real-world data from clinical documentation to research-ready phenotypes, but with varying success (Kreimeyer et al. 2017; Koch et al. 2024). Early work using NLP relied on rule-based methods, and this was followed by machine learning and deep learning approaches (Harshavardhan et al. 2025). With the advent of transformer architectures and large language models (LLMs), encoder and decoder models have set new benchmarks for information extraction (Clay et al. 2025). LLMs have successfully identified phenotypes from EHRs and comparable text (Munzir et al. 2024), including psychiatric data using LLM-assisted extraction combined with manual validation (Edwards et al. 2025) and fully automated LLM-led pipelines (Frydman-Gani et al. 2025).

Interest in AI and LLMs has prompted calls to integrate these tools into clinical psychiatric settings and healthcare research (Reddy 2024; Tian et al. 2024). However, the heterogeneity of psychiatric phenotypes makes reliable, large-scale phenotyping a formidable task, and LLMs have rarely been benchmarked on their capability to extract psychiatric symptoms from text. This gap must be addressed before LLM-based tools can move from concept to implementation. Here, we evaluate 14 open-weight LLMs for extracting psychosis-related phenotypes from free-text clinical summaries detailing core symptoms of schizophrenia. We assess whether these models can retrieve symptom presence using zero- and few-shot prompting and quantify how task performance varies across model architectures and prompting strategies.

## Methods

### Study design and sample

Participants were drawn from the Cardiff COGnition in Schizophrenia (CardiffCOGS) cohort (N=1310), a sample of individuals with severe mental illness (mainly schizophrenia, schizoaffective disorder, and bipolar disorder) recruited from South Wales and the surrounding areas. Individuals were recruited from community adult mental health services, inpatient services, and voluntary services through advertisements and mental health organisations. Participants completed a clinical research interview based on the Schedules for Clinical Assessment in Neuropsychiatry (SCAN; Wing et al. 1990). For the present study, we included only CardiffCOGS participants with clinical vignettes available. These were free-text summaries of the participants’ psychiatric secondary care notes and comprised narrative descriptions of their clinical history and presentation. Summaries were pre-prepared by trained psychiatrists or research psychologists and contained variable levels of information. Individuals were excluded from analyses if no gold-standard ratings were available for any phenotype (N=12). A small subset of participants were extracted from the dataset to use their notes for in-context few-shot learning (N=8). The final sample size used for LLM-based symptom extraction comprised 704 participants (**Figure 1**).

**Figure 1.**
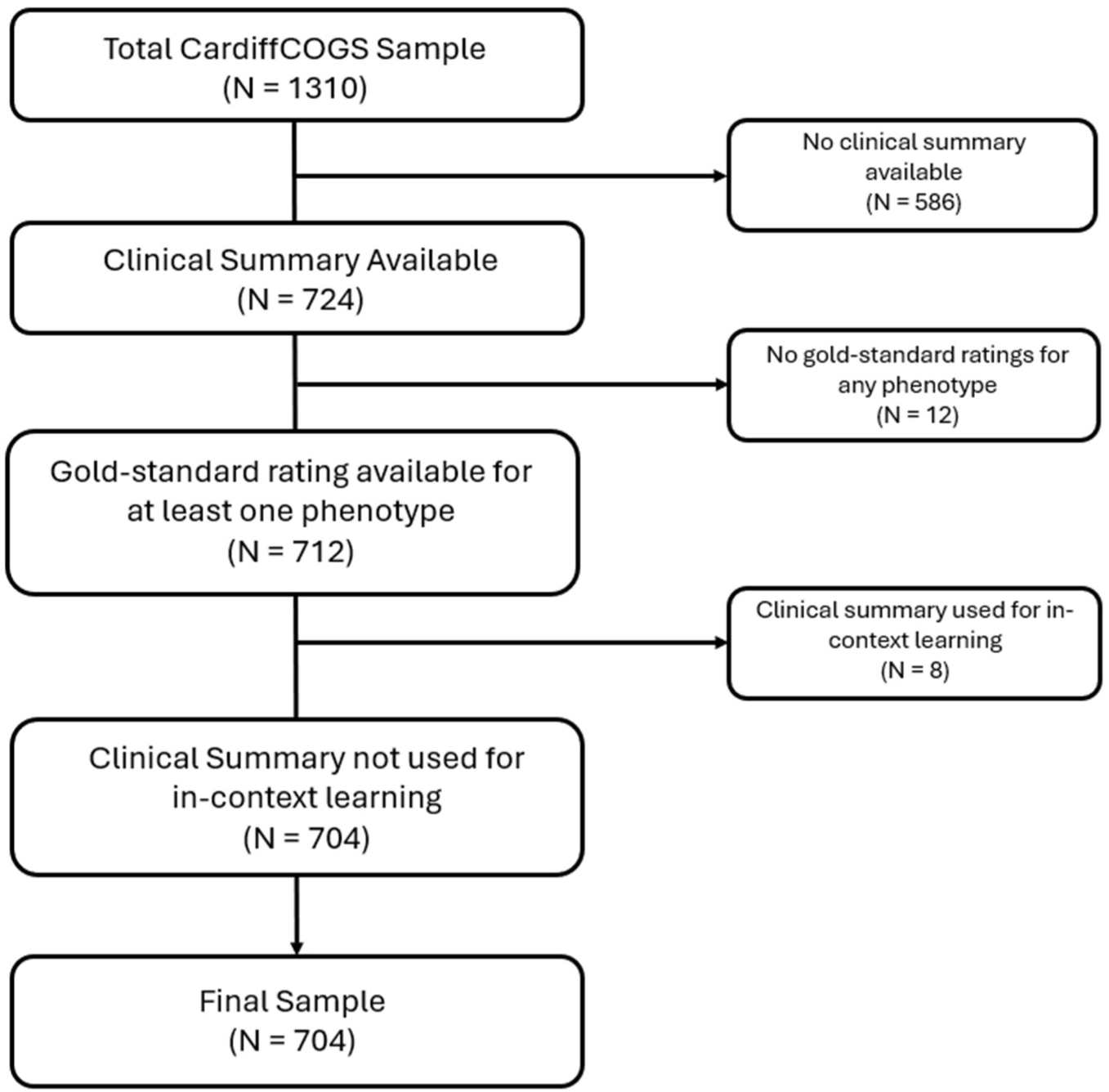
Flow chart showing ascertainment of sample size used in model evaluation.

This study follows the MLPsych Guidelines for Reporting Machine Learning Investigations in Neuropsychiatry (GREMLIN; Quinn et al. 2024; **Supplementary Materials**). CardiffCOGS received approval from the South-East Wales Research Ethics Committee (07/WSE03/110), and all participants provided written informed consent.

### Involvement of people with lived experience

Experiments were designed after engaging a panel of three people with lived experience of severe mental illness who described themselves as casual users of AI. This panel helped shape the study focus and contributed to prioritising the assessed outcomes.

### Outcomes

The eight phenotypes of interest were symptoms of schizophrenia and other psychotic disorders selected from the global symptom categories in the Scale for the Assessment of Positive Symptoms (SAPS; Andreasen 1984) and the Scale for the Assessment of Negative Symptoms (SANS; Andreasen 1989). These included hallucinations, delusions, positive formal thought disorder (PFTD), bizarre behaviour, affective flattening, alogia, anhedonia and asociality, and avolition and apathy (**Supplementary Table 1**).

In the primary analysis, we extracted a binary variable denoting symptom presence or absence from the clinical summaries. Predicted values were compared to gold-standard Global Lifetime Worst SAPS and SANS scores (Legge et al. 2021). Ratings were completed by trained staff (psychiatrists or psychology graduates) supervised by consultant psychiatrists, as previously described (Legge et al. 2021). Interrater reliability was good across SAPS and SANS items (κ = 0.72 – κ = 0.95). Binarized versions of these ratings, for comparison with LLM-based outputs, were generated as described in **Table 1**.

**Table 1.**
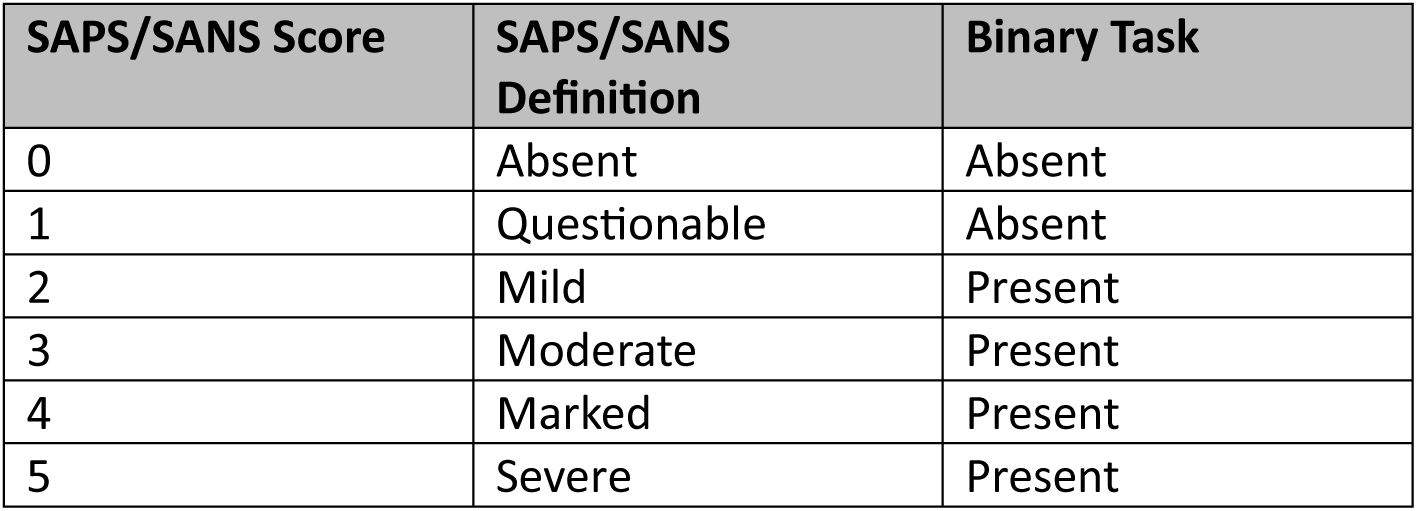
Conversion between ratings on the Scale for the Assessment of Positive Symptoms (SAPS) and Scale for the Assessment of Negative Symptoms (SANS) and the binary scale used in the present work.

| **SAPS/SANS Score** | **SAPS/SANS Definition** | **Binary Task** |
| --- | --- | --- |
| 0 | Absent | Absent |
| 1 | Questionable | Absent |
| 2 | Mild | Present |
| 3 | Moderate | Present |
| 4 | Marked | Present |
| 5 | Severe | Present |

### Model Selection and Testing

We assessed a range of LLMs as described in **Table 2**; only models with the ‘Thinking’ suffix and the Deepseek-R1 line were explicitly trained for reasoning. Clinical summaries with no personal identifiers were processed using open-source models that could be run in the secure Isambard AI Phase 2 high-performance computing (HPC) environment. Model selection was guided by past success in extracting psychiatric phenotypes (Frydman-Gani et al. 2025) and by novel releases. LLMs were not fine-tuned; however, the eight case summaries that were used to generate few-shot prompts were excluded from model evaluation for both zero- and few-shot analyses to prevent data leakage.

**Table 2.** Summary of models evaluated for phenotype extraction. LLM = Large Language Model.

| Full Model Name | Type | Developer | # Parameters |
| --- | --- | --- | --- |
| Llama-3.1-8B-Instruct | LLM | Meta | $8 \times 10^9$ |
| Llama-3.3-70B-Instruct | LLM | Meta | $7 \times 10^{10}$ |
| Llama3-OpenBioLLM-70B | LLM | Meta and aaditya | $7 \times 10^{10}$ |
| Mistral-Nemo-Instruct-2407 | LLM | Mistral AI and NVIDIA | $1.2 \times 10^{10}$ |
| Mistral-Small-24B-Instruct-2501 | LLM | Mistral AI | $2.4 \times 10^{10}$ |
| Mistral-Large-Instruct- 2411 | LLM | Mistral AI | $1.23 \times 10^{11}$ |
| gemma-3-27b-it | LLM | Google | $2.7 \times 10^{10}$ |
| medgemma-27b-text-it | LLM | Google | $2.7 \times 10^{10}$ |
| Qwen3-Next-80B-A3B-Thinking | LLM | Alibaba Cloud | $8 \times 10^{10}$ |
| Qwen3-Next-80B-A3B-Instruct | LLM | Alibaba Cloud | $8 \times 10^{10}$ |
| Qwen3-235B-A22B-Instruct-2507 | LLM | Alibaba Cloud | $2.35 \times 10^{11}$ |
| Qwen3-235B-A22B-Thinking-2507-FP8 | LLM | Alibaba Cloud | $2.35 \times 10^{11}$ |
| DeepSeek-R1-Distill-Qwen-32B | LLM | DeepSeek and Alibaba Cloud | $3.3 \times 10^{10}$ |
| DeepSeek-R1-Distill-Llama-70B | LLM | DeepSeek and Meta | $7.1 \times 10^{10}$ |
| mental-bert-large-uncased | Encoder | Google and AI for Mental Health | $3.4 \times 10^8$ |
| mental-longformer-base-4096 | Encoder | AllenAI and AI for Mental Health | $1.49 \times 10^8$ |
| google_bigbird-roberta-large | Encoder | Google | $3.55 \times 10^8$ |
| mental_mental-bert-base-uncased | Encoder | Google and AI for Mental Health | $1.1 \times 10^8$ |
| gatortron-base | Encoder | University of Florida NLP Group | $3.45 \times 10^8$ |

LLMs were run using Python v3.11.7 (Van Rossum and Drake 2009). For efficiency, we used *vLLM* (Kwon et al. 2023) and loaded models using *bitsandbytes* (Dettmers et al. 2022a; Dettmers et al. 2022b) for in-flight 4-bit quantisation, or if available, pre-quantised models were loaded in 8-bit. Where available, sampling parameters followed recommendations in the HuggingFace model card; where none were provided, default settings were used with the max token length set to 32,768. Inference was run three times per model, with mean evaluation metrics reported. Scripts will be made available upon publication at https://github.com/locksk/llm-scz-symptoms.

For comparison, we also assessed several fine-tuned encoder-only models, most of which had been pre-trained on clinical text and were based on BERT or BERT-like architectures. Briefly, we split our sample into training, validation, and test sets following a 7:2:1 ratio. Following the LLM analyses, we trained each model to predict each symptom from the clinical summaries using a binary classifier, optimising hyperparameters with Optuna (Akiba et al. 2019). Model performance was evaluated solely on the test set. Full details are provided in the **Supplementary Methods**.

### Prompts

We tested both zero-shot and few-shot prompting strategies. Prompts were developed on a subsample of the CardiffCOGS clinical summaries (N = 50) using *Llama-3.3-70B-Instruct* as the reference model. Prompts were evaluated under zero- and few-shot conditions, with final prompts selected based on performance across the greatest number of symptoms. For few-shot prompts, examples of symptom presence and absence were selected from clinical vignettes in the DRAGON-Data harmonised resource (Lynham et al. 2023). Symptom definitions used in prompts are provided in **Supplementary Table 1**.

A total of 5,632 zero-shot prompts (704 participants x 8 symptoms) were fed to each model as described in **Supplementary Table 2**. In the few-shot condition, four brief examples were provided for each symptom alongside the gold-standard rating. Examples were presented as text excerpts containing both present and absent cases, as opposed to complete case summaries. All prompts supplied to the LLMs followed the “system” and “user” format, except for the DeepSeek R1 family of models. In those, all information was included within the “user” prompt, and reasoning was enforced via addition of “<THINK>\n” before the output, as recommended in the model documentation (DeepSeek-AI 2025). The HuggingFace *apply_chat_template* method was used to format prompts for each model.

### Evaluation

Data were analysed in R v4.4.1 via the RStudio IDE (R Core Team 2021). Model output was cleaned where additional text accompanied the desired answer, as commonly seen in reasoning model outputs. Metrics were calculated at the per-symptom level, as described in **Supplementary Table 3**. As symptoms can be subject to class imbalance, we included measures that are robust to this issue in our evaluation (e.g., F1, Balanced Accuracy). Models were ranked according to mean macro-average F1 score across symptoms, which determined which LLMs were taken forward for downstream analysis. F1 scores can range from 0 to 1, where scores closer to 1 represent better model performance; unlike accuracy, F1 does not reward correctly predicting the majority negative class, making it better suited to imbalanced labels.

To validate LMM predictions, we regressed each of the eight extracted phenotypes from the top-performing model against clinical variables known to be associated to the severity of schizophrenia symptoms (Legge et al. 2020; Cardno et al. 2025). These variables included age at onset, course of disorder, ever prescribed clozapine, ever prescribed depot medications, and lifetime worst scores on the Global Assessment Scale (Endicott et al. 1976). All logistic regression models included sex, age at interview, and schizophrenia diagnosis as covariates. The False Discovery Rate (FDR; Benjamini and Hochberg 1995) was used to correct for multiple comparisons.

Separately, we used ordinal regression to test whether demographic variables (i.e., age, sex, diagnosis, self-reported ethnicity) or text features (i.e., number of characters in the case summary, summariser group; see **Supplementary Note**) were associated with the number of symptoms correctly predicted by the top-performing LLM. Additionally, and separately for each symptom, we sampled 20 participants where the LLM-based phenotype did not match the gold-standard annotation. Case notes from these participants were used to perform a qualitative error analysis to identify broad error categories occurring in our dataset.

Finally, selecting three comparable models of different sizes, we tested additional prompts to assess whether few-shot prompting improved performance. Where task performance comparisons were required, McNemar’s test was used to compare misclassification rates, with a False Discovery Rate (FDR) correction.

## Results

### Sample Characteristics

We included 704 participants, of whom over 90% had a diagnosis of psychosis (**Table 3**). Clinical summaries were written by 17 researchers; in some instances, multiple summarises contributed to a single case note. The distribution of gold-standard ratings for the eight global symptoms is shown in **Supplementary Figure 1**.

**Table 3.** Demographic and clinical variables for the total sample. Mean (SD) is reported for continuous variables, and n (%) is reported for categorical variables.

| **Variable** | **Mean (SD); n (%)** |
| --- | --- |
| Sex |  |
| Male | 425(60%) |
| Female | 279 (40%) |
| Age | 42.8 (11.9) |
| Ethnic Background |  |
| White | 676 (96%) |
| Black, Black British, Caribbean or African | 4 (0.6%) |
| Asian or Asian British | 10 (1.4%) |
| Mixed or multiple ethnic groups | 13 (1.9%) |
| Unknown | 1 |
| Diagnosis |  |
| Schizophrenia | 384 (55%) |
| Schizoaffective Depressed | 81 (12%) |
| Schizoaffective Bipolar | 60 (8.5%) |
| Other psychotic | 122 (17%) |
| Bipolar 1 | 16 (2.3%) |
| Bipolar 2 | 5 (0.7%) |
| Depression | 28 (4%) |
| Other | 6 (0.9%) |
| Unknown | 2 |
| Number of Admissions | 4.5 (5.2) |
| Unknown | 11 |

### Assessing Model Performance

No model demonstrated optimal performance across all eight symptoms (**Table 4**). Maximum macro-averaged F1 scores were attained primarily by Qwen LLMs, though other models performed at comparable levels (**Figure 2**). A beta generalised linear mixed effect model demonstrated that positive symptoms had higher macro-average F1 scores than negative symptoms (β = 0.467, SE = 0.027, *p* = 1.32×10^-66^). The best performance was for Positive Formal Thought Disorder (PFTD), followed by delusions, and hallucinations, which both demonstrated more consistent F1 scores across models. Negative symptoms of anhedonia and asociality, as well as avolition and apathy, had the poorest performance with no model reaching a macro-average F1 score above 0.6. *Qwen3-235B-A22B-Thinking-2507-FP8* was the top performing model with mean macro-averaged F1 score of 0.648 (SD = 0.072; range = 0.553 – 0.793) across symptoms (**Table 5**). Finally, despite many being pre-trained specifically on clinical text, encoder models were outperformed by LLMs in most tasks. Full results are in **Supplementary Figure 2** and **Supplementary Tables 4 – 21**.

**Table 4.** Best per-symptom prediction performance based on Macro-Average F1 Score. PFTD = Positive Formal Thought Disorder.

| Symptom | Domain | Macro-Average F1 Score | F1 Score (Present) | Best Model |
| --- | --- | --- | --- | --- |
| Hallucinations | Positive | 0.684 | 0.853 | Qwen 3 A235B Instruct |
| Delusions | Positive | 0.725 | 0.931 | Qwen 3 A235B Thinking |
| PFTD | Positive | 0.820 | 0.770 | Qwen 3 A235B Instruct |
| Bizarre Behaviour | Positive | 0.666 | 0.913 | Qwen 3 Next Instruct |
| Affective Flattening | Negative | 0.627 | 0.591 | Qwen 3 A235B Thinking |
| Alogia | Negative | 0.638 | 0.584 | Gemma-3-27B |
| Anhedonia and Asociality | Negative | 0.575 | 0.719 | Qwen 3 Next Instruct |
| Avolition and Apathy | Negative | 0.576 | 0.740 | Qwen 3 A235B Thinking |

**Figure 2.**
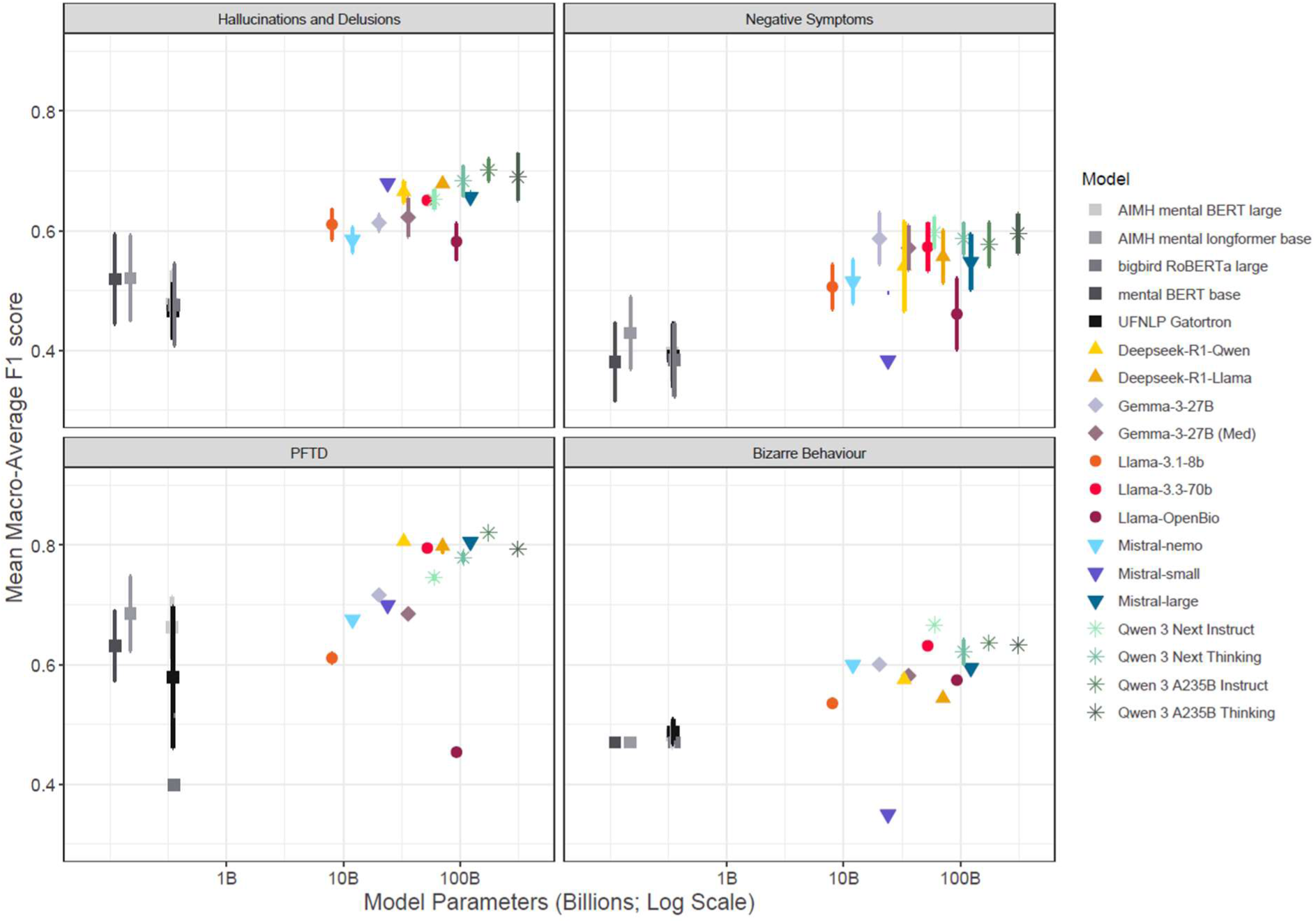
Comparison of Mean Macro-Average F1 Score across different symptoms and models in a zero-shot setting. Model size, indexed by number of parameters, is represented on the log scale on the x-axis. Results for clinically similar features have been pooled. Error bars represent standard deviation around the mean.

**Table 5.** Per-model average evaluation metrics across symptoms. Models are ordered by their mean macro-averaged F1 score. Balanced accuracies are also reported. Models with an asterisk (*) are those that are prioritised for later few-shot analyses.

| Rank | Model | Mean Macro-F1 (SD) | Median Macro-F1 (MAD) | Mean BAC (SD) | Median BAC (MAD) |
| --- | --- | --- | --- | --- | --- |
| 1 | Qwen 3 A235B Thinking* | 0.647 (0.072) | 0.63 (0.066) | 0.071 (0.67) | 0.639 (0.051) |
| 2 | Qwen 3 A235B Instruct | 0.645 (0.087) | 0.628 (0.097) | 0.073 (0.663) | 0.65 (0.066) |
| 3 | Qwen 3 Next Thinking* | 0.638 (0.069) | 0.617 (0.073) | 0.074 (0.659) | 0.637 (0.074) |
| 4 | Qwen 3 Next Instruct | 0.635 (0.052) | 0.631 (0.052) | 0.052 (0.647) | 0.626 (0.019) |
| 5 | Llama-3.3-70b | 0.624 (0.075) | 0.623 (0.042) | 0.067 (0.638) | 0.627 (0.035) |
| 6 | Deepseek-R1-Llama | 0.616 (0.091) | 0.596 (0.108) | 0.067 (0.653) | 0.639 (0.063) |
| 7 | Mistral-large | 0.611 (0.089) | 0.6 (0.08) | 0.08 (0.643) | 0.627 (0.053) |
| 8 | Gemma-3-27B | 0.609 (0.049) | 0.607 (0.028) | 0.05 (0.632) | 0.614 (0.019) |
| 9 | Deepseek-R1-Qwen* | 0.608 (0.102) | 0.61 (0.092) | 0.077 (0.649) | 0.639 (0.069) |
| 10 | Gemma-3-27B (Med) | 0.599 (0.047) | 0.598 (0.038) | 0.048 (0.622) | 0.609 (0.03) |
| 11 | Mistral-nemo | 0.56 (0.06) | 0.559 (0.062) | 0.051 (0.585) | 0.571 (0.021) |
| 12 | Llama-3.1-8b | 0.551 (0.057) | 0.549 (0.086) | 0.044 (0.574) | 0.581 (0.058) |
| 13 | Llama-OpenBio | 0.508 (0.074) | 0.502 (0.075) | 0.033 (0.549) | 0.545 (0.038) |
| 14 | Mistral-small | 0.5 (0.175) | 0.489 (0.274) | 0.089 (0.618) | 0.605 (0.111) |

### Validation Analysis

An illustration of all of our validation analysis results is given in **Figure 3**. Poorer functioning on the Global Assessment Scale was associated with *Qwen3-235B-A22B-Thinking-2507-FP8* outputs for hallucinations (OR = 0.938, 95% CI = 0.912 - 0.965, *p* = 2.78 × 10^-5^), delusions (OR = 0.922, 95% CI = 0.894 - 0.95, *p* = 1.15 × 10^-6^), positive formal thought disorder (OR = 0.930, 95% CI = 0.908 - 0.951, *p* = 6.74 × 10^-9^), bizarre behaviour (OR = 0.942, 95% CI = 0.918 - 0.965, *p* = 1.95 × 10^-5^), affective flattening (OR = 0.960, 95% CI = 0.939 - 0.981, *p* = 0.002), and alogia (OR = 0.966, 95% CI = 0.945 - 0.987*, p* = 0.016). A more severe disorder course was also associated with the presence of hallucinations (OR = 1.947, 95% CI = 1.535 - 2.484, *p* = 4.27 × 10^-5^) and affective flattening (OR = 1.276, 95% CI = 1.071 - 1.525, *p* = 0.027).

**Figure 3.**
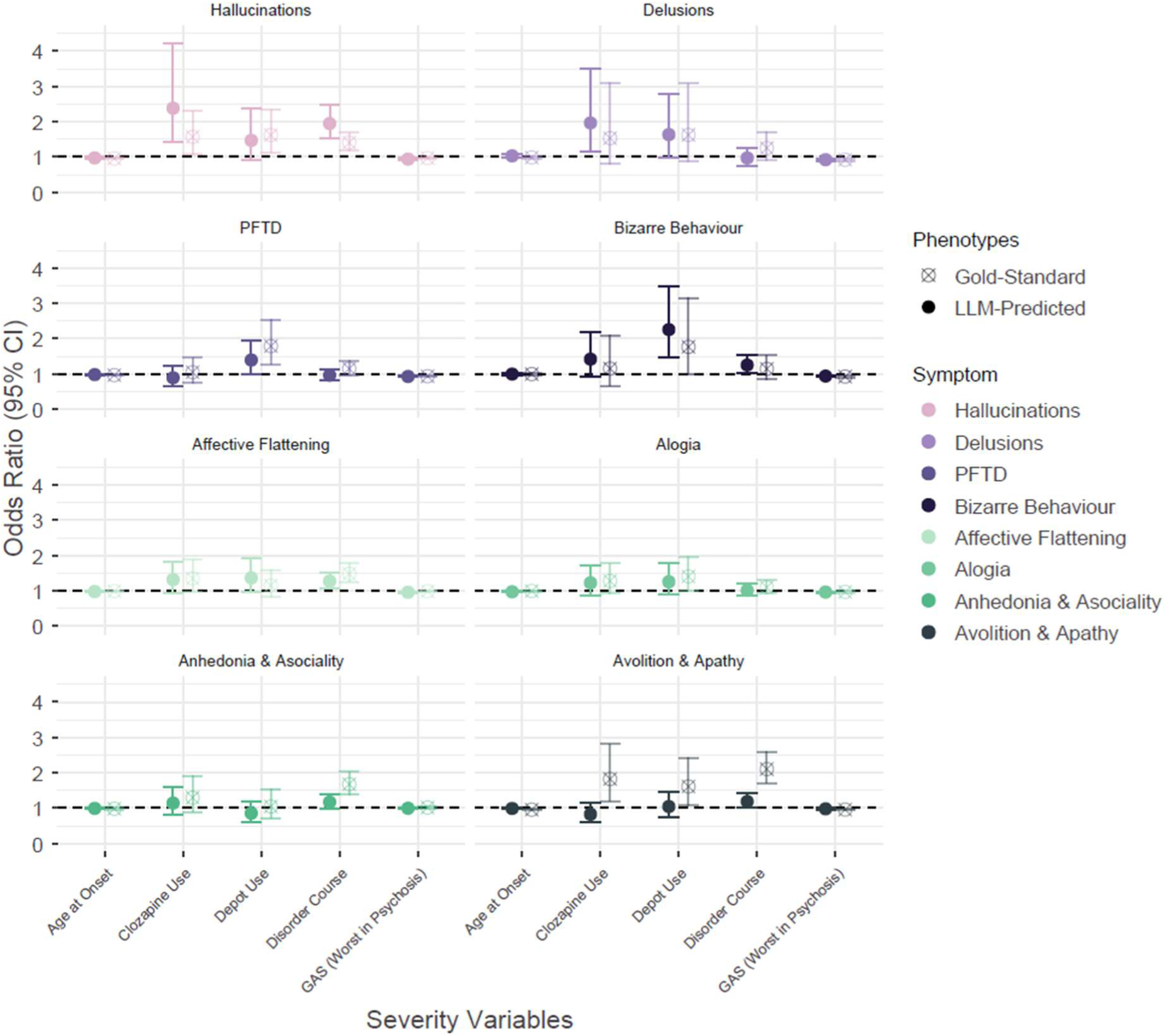
Results from validation analysis showing associations between LLM-predicted symptoms and variables relating to schizophrenia severity. All results are from univariate Generalised Linear Models controlling for diagnosis, age and sex. For comparison, associations between gold-standard symptoms with severity variables are also provided.

Symptoms predicted by *Qwen3-235B-A22B-Thinking-2507-FP8* were also associated with medication use. Lifetime depot prescription was associated with bizarre behaviour (OR = 2.257, 95% CI = 1.474 - 3.487, *p* = 8.25 × 10^-4^). Lifetime clozapine use was associated with hallucinations (OR = 2.384, 95% CI = 1.409 - 4.206, *p* = 0.005) and delusions (OR = 1.963, 95% CI = 1.144 - 3.501, *p* = 0.035). Full results for both LLM-predicted and gold-standard symptom phenotypes are in **Supplementary Tables 22 – 25.**

### Error Analysis

We analysed errors from *Qwen3-235B-A22B-Thinking-2507-FP8,* the highest-performing model (mean macro-average F1 score = 0.648). The median number of correctly predicted symptoms was six out of eight, and over 80% of participants had 5 or more symptoms correctly predicted. Log-transformed case summary length was associated with the number of correct symptom predictions (β = 0.113, SE = 0.023, *p* = 1×10⁻⁶), accounting for 11% of variance in task success. Full results are in **Supplementary Table 26.**

At a per-symptom level, false negatives were more common than false positives, except for hallucinations and PFTD (**Figure 4**). We examined a subset of 20 errors for each symptom, using the thinking tokens (i.e., verbalisations of the model’s reasoning process) to identify the main reasons underlying incorrect predictions. We identified six broad classes of error: hypersensitivity, hyposensitivity, missing information, conflicting information, symptom misattribution, and definition errors (See **Figure 5**; full results in **Supplementary Table 27**). We did not identify any instances of seemingly fabricated responses or confabulations.

**Figure 4.**
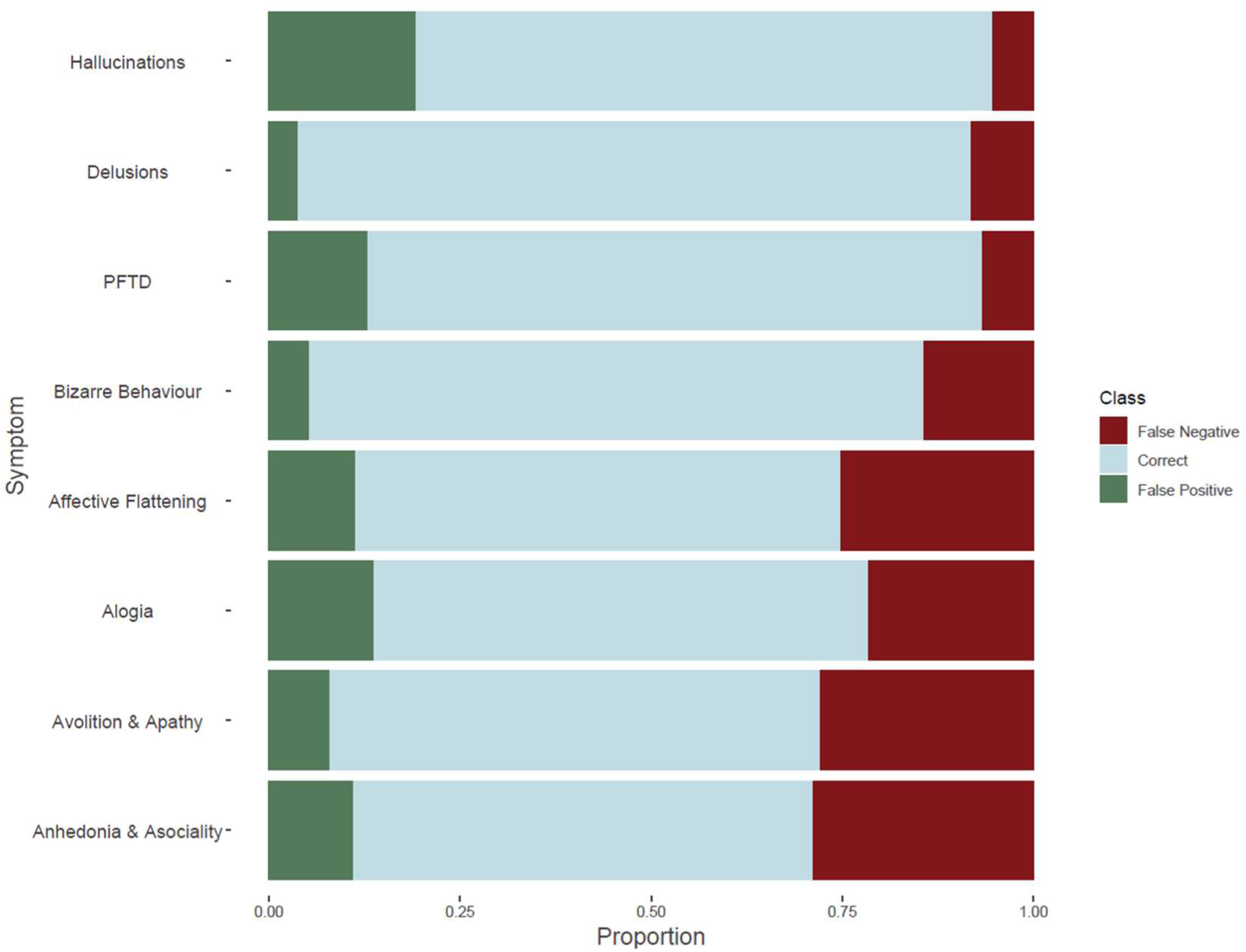
Correct ratings and error rates across the 8 symptoms. Bars show the proportion of predictions made by the top model (Qwen-3-235B) that were correct or incorrect, alongside showing the breakdown of false positives and negative amongst the errors.

**Figure 5.**
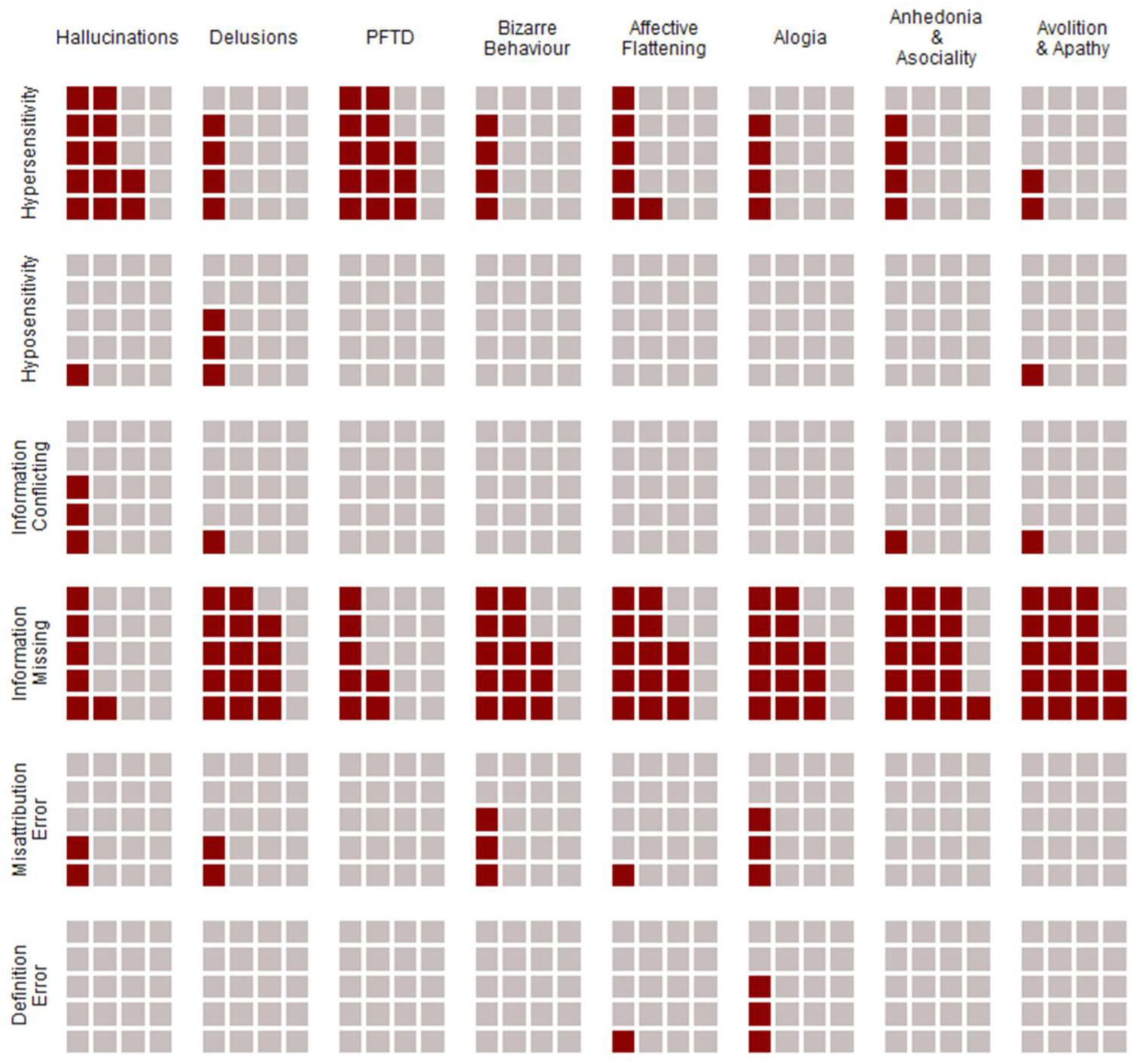
Types of Errors made by LLMs in Symptom Prediction. For each symptom, 20 errors were randomly sampled, and the types of errors categorised. Red blocks represent the number of errors of each type made per symptom.

Missing information was one common error type. Gold standard ratings were based on clinical interviews and case notes, whereas LLM predictions relied only on case note summaries. This information asymmetry implies models lacked, at least sometimes, the context needed to make a correct rating, leading to an excess of false negatives. To investigate this further, we identified false negatives (391 symptom ratings from 308 participants) and reclassified them using only case note summaries (**Supplementary Table 28**). Reclassification was performed by trained raters (LME, SEL), with high interrater reliability (κ^ = 0.881). When symptoms were assessed solely on case note summaries, nearly half (42.5%) of ratings changed. Furthermore, model performance improved significantly when benchmarked against reclassified ratings (χ^2^_McNemar_ = 50.919, *p* = 1×10⁻¹²). Based on the Miettinen-Nurminen confidence intervals (Fagerland et al. 2015), the proportion of reclassification was significantly different between the positive symptoms of hallucinations and delusions *versus* the negative symptoms, with roughly 35% more instances resulting in reclassification in the former than the latter (*p* = 2×10⁻^9^).

### Zero-Shot versus Few-Shot

We observed no improvement from using Few-Shot prompting across Deepseek-Qwen-32B (χ^2^_McNemar_ = 0.344, *p* = 0.707), Qwen3-Next-80B (χ^2^ = 2.450, *p* = 0.354), and Qwen3-235B (χ^2^_McNemar_ = 0.141, *p* = 0.707). Full evaluation metrics from the few-shot analysis are shown in **Supplemental Tables 29 – 37**, and per-model per-symptom McNemar’s test results in **Supplementary Table 38.**

### Energy Use Calculations

Energy use in kilowatt hours (*kWh*) was converted to CO_2_ emissions by multiplying by the carbon intensity of the UK grid at 233 gCO2/kWh. Power consumption was estimated at 660W, in line with Isambard AI caps for each NVIDIA GH200 Grace Hopper Superchip. We used 37.68 node hours, equivalent to 150.72 GPU hours, to perform all LLM analyses, equating to 100 kWh or 23.3 kg CO_2_.

## Discussion

Here, we test the ability of Large Language Models to extract information about symptoms of schizophrenia and other psychotic disorders from summaries of clinical notes. We demonstrate variable extraction performance based on the target symptom, the LLM used, and the length of the case note. No models attained macro-average F1 scores greater than 0.9 across any symptom, plateauing well below F1 scores suggestive of potential clinical or research utility. This echoes findings from research using encoder models (Zeinali et al. 2024), where physical symptoms were extracted more accurately than psychiatric symptoms. Indeed, our results suggest that general-purpose language models (both encoders and LLMs) struggle with psychiatric writing, regardless of pre-training source or parameter scale.

### Individual LLM performance

Qwen models achieved the highest performance both across most individual symptoms and on average, consistent with their position among the largest models tested exceeding 80 billion parameters. For certain symptoms, smaller models outperformed similarly sized alternatives, for example *Mistral-Small-24B* for hallucinations and delusions and *DeepSeek-Qwen-32B* for PFTD (**Figure 2**). Fine-tuned encoders were generally outperformed by LLMs, despite encoders being pre-trained on clinical documentation, scientific text, or mental health corpora. This contrasts with findings that fine-tuned encoders out-perform LLMs on symptom detection in emergency department records, partly due to improved negation handling (Diaz Ochoa et al. 2025). Our results are symptom dependent and, for zero-shot analyses, are broadly comparable to those from a study of Spanish-language EHRs for equivalent symptoms (Frydman-Gani et al. 2025). While several LLMs assessed here were also used in that study, performance differences between the two studies at the model and symptom levels suggest these information extraction tasks might be highly sensitive to the input dataset, at least as currently implemented. This variability can be expected when using general purpose LLMs or when adapting them to specialised tasks through in-context learning alone, which supports the exploration of fine-tuning and other task-specific training approaches as future avenues of work (Bucher and Martini 2024).

### Positive and Negative Symptoms

PFTD was the best extracted symptom, with several models attaining macro-averaged F1 scores at or above 0.8. Positive (hallucinations and delusions) were better extracted than negative symptoms (affective flattening, alogia, anhedonia and asociality, and avolition and apathy). Furthermore, LLM performance for hallucinations and delusions was fairly consistent across models and was generally characterised by high precision and recall with lower specificity. This suggests models applied to our cohort are better at identifying the presence of symptoms that track additional behaviours (i.e., positive symptoms) rather than those which relate to impairments or restrictions (i.e., negative symptoms). No consistent pattern emerged in precision, recall, and specificity across models for other symptoms.

Among negative symptoms, performance for anhedonia and asociality, as well as avolition and apathy, was poor, with no model exceeding a macro-F1 score of 0.580. Negative symptoms represent an absence of typical behaviour, so describing them relies on negation and gradation structures: for example, “*lack of speech*” would signal alogia, “*reduced affective range*” could be used for affective flattening, or “*does not seek social contact*” for anhedonia. Negations in particular have been a historical challenge for NLP that LLMs have not fully solved (Varshney et al. 2025). As such, performance for these symptoms may be limited, in part, by the linguistic features that define them. Negative symptoms are important features of schizophrenia and are known to predict poor functional outcomes (Pogue-Geile 1989; Fervaha et al. 2014; Galderisi et al. 2018). They also respond little to standard antipsychotics (Correll and Schooler 2020), which could result in them being underreported in comparison to positive symptoms. This notion is supported by the error analysis in which a lack of information in the case summary was observed for the majority of incorrect classifications of negative symptoms made by the model. If these constraints affect how negative symptoms are described in clinical notes, they would reinforce the performance gap we observe in how positive and negative symptoms are extracted by LLMs, reifying a bias between these two symptom domains.

### Types of Error

Exploring the results of our top-ranked LLM showed that associations between clinical variables and LMM-predicted symptoms closely resembled those with gold-standard annotations, despite macro-F1 scores not suggestive of potential clinical utility. Across 64 pairings of symptoms with clinical variables, effect sizes from the two sets of phenotypes correlated at r = 0.68 and agreed in direction for 79.7% of pairs (**Supplementary Figure 3**). However, effect sizes involving positive symptoms also had similar magnitudes in both the LLM-predicted and gold-standard analyses, while the effects involving LLM-predicted negative symptoms were more likely to shrink towards the null (**Supplementary Table 22 - 25**). Our interpretation of these observations is that LLM errors are not simply randomly distributed. Misclassification and measurement error result in a systematic attenuation of the associations between noisy variables and their covariates, at least when samples are adequately powered to detect the effects (Neuhaus 1999; Liu and Zhang 2017; Hamra 2022). We do not see this attenuation in positive symptoms, where 80% of associations surviving FDR correction in the gold-standard data were recovered to at least nominal significance in the LLM-derived data. The error analysis highlights that misclassifications in this domain were partly driven by model hypersensitivity. While incorrect, these ratings are not necessarily uninformative and may reflect features correlated with symptoms such as general psychopathology or sub-clinical/prodromal phenomena.

The associations between negative symptoms and the clinical variables were attenuated, with only 2 out of 6 FDR-significant gold-standard associations being recovered in the LLM-based analysis, consistent with potential misclassification. It is noteworthy that symptoms from this domain were particularly enriched for false negatives and missing information errors (**Figure 4**, **Figure 5**). Re-examination of false negatives by trained raters concluded that under half of them would be defensible from the content of the clinical summaries, supporting that the LLM-based inference was valid given the available free text. We argue that missing information in the free text may be a driver of non-differential misclassification, leading to reduced effective sample size and thus power to detect true effects (Kestenbaum 2009).

### Practical Implications

A practical implication of our study is that LLM-predicted phenotypes could be considered for performing first-pass or hypothesis-generating analyses in large cohorts, provided their interpretation is framed in the context of probable measurement error. However, it should be noted the present work still identifies clear discrepancies between clinician-standard and LLM-predicted ratings, particularly for negative symptoms. Those, alongside biases identified elsewhere in the wider clinical NLP literature (Cross et al. 2024), remain a clear limitation of LLMs and other information retrieval methodologies. Finally, we report that case summary length predicted the number of correctly extracted symptoms. This was expected as longer case summary should be more likely to contain more symptom-relevant text. However, many models have showed declining performance with increasing input length (Levy et al. 2024), and even frontier LLMs with large context windows struggle to retrieve information from the middle of long prompts (Janik 2024; Salvatore et al. 2025). Given our analysis used clinical summaries instead of full case notes, the association between summary length and accuracy should be probed in future experiments with increased context windows, as it could attenuate or reverse if complete EHRs are used as input.

### Limitations

We used summaries of psychiatric case notes rather than full EHRs, which limits our conclusions to that type of input text. Our results may therefore not generalise to EHR-based work, though it should be noted that both data types are affected by similar limitations. Second, the error analysis confirmed that incomplete records remain a bottleneck for LLMs in this task, particularly for negative symptoms. Missing data is a known problem in psychiatric EHRs (Madden et al. 2016), arising from difficulties in coordinating care across healthcare systems, from limited or absent documentation standards, and from clinician uncertainty about what constitutes relevant information for a given case (Chivilgina et al. 2022). Patient history and clinician expertise likely contribute to systematic variation in how symptoms are recorded, potentially resulting in incomplete clinical pictures that, for example, may only include symptoms directly related to a current diagnosis. Discordance between EHRs and patient self-report has also been demonstrated (Pakhomov et al. 2008), and may be greater in mental health where stigma and limited insight can widen the gap between clinician- and patient-reported symptoms (Aaslestad and Skuggevik 2018). As case notes and electronic records are increasingly used as the basis for clinical and research tools, the adoption or strengthening of best-practice recommendations for clinical documentation could help to address these challenges.

### Future Directions

The capabilities of general-purpose LLMs for information extraction and natural language understanding are advancing rapidly. Although our results do not support that they can yet extract information on psychotic symptoms from psychiatric notes to the standard needed for clinical use, the gap between LLM-derived and clinician-derived phenotypes is likely to narrow. There are several areas that must be addressed for such a gap to become small enough to warrant the effort and investment required for implementation trials.

First, our evaluation targeted global SAPS and SANS symptom categories. Future work should address constituent sub-symptoms, such as the subdivision of “delusions”, for example into persecutory, grandiose, somatic, and passivity variants. Work should also move from merely identifying whether symptoms are present to quantifying their severity. The use of NLP to extract severity metrics has been understudied (Koleck et al. 2019); automating this would allow a richer characterisation of individual experiences and support monitoring of symptom changes both over time and with treatment.

Second, performance was consistently poorer for negative than for positive symptoms. While this was partly expected due to the poorer handling of negations by LLMs, negative symptoms are also intrinsically more difficult to identify (e.g., “affective flattening” refers specifically to the expression of emotions, not their subjective experience) and this is likely to be reflected in input texts. Contrary to expectations from prior work (Frydman-Gani et al. 2025), in-context learning via few-shot prompting provided no benefit here. Alternative approaches might include combining clinical records with multimodal data sources (e.g., audio transcripts, open-ended writing, neuroimaging data, omics, etc), increasing the available information and potentially addressing some of the pitfalls identified in the error analysis (Wright et al. 2026).

Third, we considered fine-tuning of LLMs as out of scope for this study, but it is another candidate solution to improve task performance. However, it must be weighed against computational demands and concerns about clinical data governance and AI-oriented uses. In a study using Spanish-language EHRs, fine-tuning was shown to enable small-scale LLMs to identify psychiatric phenotypes with an accuracy comparable to much larger untuned models (Frydman-Gani et al. 2025). Accurate models with small parameter sizes could reduce the computational resources required to run experiments similar to ours, making them transferrable across clinical and research contexts even when large-scale GPU-accelerated HPC systems are not available.

## Conclusions

Extracting accurate symptom information from electronic health records is a necessary step towards the high-throughput deep phenotyping of individuals with psychiatric disorders. For schizophrenia and other psychotic disorders, this is not yet feasible at the standard required for clinical use using general-purpose LLMs. Performance varies with the model, the target symptom, and the case note length, with positive symptoms consistently better extracted than negative symptoms. Few-shot prompting provided no benefit, suggesting researchers should look beyond in-context learning and towards supervised, or parameter-efficient fine-tuning as a way to increase task performance in LLMs. We also note that successful extraction of schizophrenia symptoms may be limited by the existing sources of clinical text. This is especially the case when they do not contain a complete, accurate, and accessible account of a patient’s health and care. For example, in our analysis we found some discrepancies between what was indicated in the clinical notes and the actual symptom recorded for the participant, due to the fact that symptom ratings considered multiple sources that were not available for this analysis. In this respect, it is not just the underlying technology of LLMs that must be improved, but also the manner in which unstructured health data is recorded both within, and between patients, clinicians, and service providers. Nevertheless, even with current limitations, both LLM-extracted and gold-standard symptoms had similar associations with clinical variables. This supports that general-purpose LLMs may already be adequate for conducting preliminary analyses of large-scale psychiatric test datasets, informing further experimental design while time-consuming tasks such as manual reviews are taking place in parallel.

## Data Availability

Code for running all parts of the analysis is available online at https://github.com/locksk/llm-scz-symptoms. To comply with the ethical and regulatory framework of the CardiffCOGS cohort, access to individual-level data requires a collaboration agreement with Cardiff University. Requests to access deidentified datasets, data dictionaries, and other summaries from the CardiffCOGS cohort should be directed to Professor James Walters.

## References

1. Aaslestad, P., Bakke, M.C.A., Ringen, P.A. and Hem, E. 2026. The patient’s voice and the writer’s role in mental health records. Tidsskrift for Den norske legeforening. Available at: https://tidsskriftet.no/en/2026/03/original-article/patients-voice-and-writers-role-mental-health-records [Accessed: 23 June 2026].

2. Aaslestad, P. and Skuggevik, E. 2018. The patient as text: the role of the narrator in psychiatric notes, 1890-1990. CRC Press.

3. Aboraya, A. et al. 2018. Measurement-based Care in Psychiatry-Past, Present, and Future. Innovations in Clinical Neuroscience 15(11–12), pp. 13–26.

4. Akiba, T., Sano, S., Yanase, T., Ohta, T. and Koyama, M. 2019. Optuna: A Next-Generation Hyperparameter Optimization Framework. In: The 25th ACM SIGKDD International Conference on Knowledge Discovery & Data Mining. pp. 2623–2631.

5. Andreasen, N.C. 1984. The Scale for the Assessment of Positive Symptoms. Iowa City, IA: University of Iowa.

6. Andreasen, N.C. 1989. The Scale for the Assessment of Negative Symptoms (SANS): conceptual and theoretical foundations. The British journal of psychiatry 155(S7), pp. 49–52.

7. Arango, C. and Carpenter, W.T. 2010. The Schizophrenia Construct: Symptomatic Presentation. In: Schizophrenia. John Wiley & Sons, Ltd, pp. 09–23. Available at: https://onlinelibrary.wiley.com/doi/abs/10.1002/9781444327298.ch2 [Accessed: 23 February 2026].

8. Benjamini, Y. and Hochberg, Y. 1995. Controlling the False Discovery Rate: A Practical and Powerful Approach to Multiple Testing. Journal of the Royal Statistical Society: Series B (Methodological) 57(1), pp. 289–300. doi: 10.1111/j.2517-6161.1995.tb02031.x.

9. Bucher, M.J.J. and Martini, M. 2024. Fine-Tuned ‘Small’ LLMs (Still) Significantly Outperform Zero-Shot Generative AI Models in Text Classification. Available at: https://arxiv.org/abs/2406.08660v2 [Accessed: 4 June 2026].

10. Cardno, A.G. et al. 2025. Associations of psychotic symptom dimensions with clinical and developmental variables in twin and general clinical samples. The British Journal of Psychiatry 226(1), pp. 16–23. doi: 10.1192/bjp.2024.129.

11. Chivilgina, O., Elger, B.S., Benichou, M.M. and Jotterand, F. 2022. “What’s the best way to document information concerning psychiatric patients? I just don’t know”—A qualitative study about recording psychiatric patients notes in the era of electronic health records. PLOS ONE 17(3), p. e0264255. doi: 10.1371/journal.pone.0264255.

12. Clay, B., Bergman, H.I., Salim, S., Pergola, G., Shalhoub, J. and Davies, A.H. 2025. Natural language processing techniques applied to the electronic health record in clinical research and practice - an introduction to methodologies. Computers in Biology and Medicine 188, p. 109808. doi: 10.1016/j.compbiomed.2025.109808.

13. Correll, C.U. and Schooler, N.R. 2020. Negative Symptoms in Schizophrenia: A Review and Clinical Guide for Recognition, Assessment, and Treatment. Neuropsychiatric Disease and Treatment 16, pp. 519–534. doi: 10.2147/NDT.S225643.

14. Cross, J.L., Choma, M.A. and Onofrey, J.A. 2024. Bias in medical AI: Implications for clinical decision-making. PLOS Digital Health 3(11), p. e0000651. doi: 10.1371/journal.pdig.0000651.

15. DeepSeek-AI. 2025. DeepSeek-R1: Incentivizing Reasoning Capability in LLMs via Reinforcement Learning. Available at: https://arxiv.org/abs/2501.12948.

16. Dettmers, T., Lewis, M., Belkada, Y. and Zettlemoyer, L. 2022a. LLM.int8(): 8-bit Matrix Multiplication for Transformers at Scale. *arXiv preprint arXiv:2208.07339*.

17. Dettmers, T., Lewis, M., Shleifer, S. and Zettlemoyer, L. 2022b. 8-bit Optimizers via Block-wise Quantization. 9th International Conference on Learning Representations, ICLR.

18. Edwards, A., Pardiñas, A.F., Kirov, G., Rees, E. and Camacho-Collados, J. 2025. Large Language Model–Supported Identification of Intellectual Disabilities in Clinical Free-Text Summaries: Mixed Methods Study. JMIR AI 4(1), p. e72256. doi: 10.2196/72256.

19. Edwards, P.J., Roberts, I., Clarke, M.J., DiGuiseppi, C., Woolf, B. and Perkins, C. 2023. Methods to increase response to postal and electronic questionnaires. Cochrane database of systematic reviews (11).

20. Endicott, J., Spitzer, R.L., Fleiss, J.L. and Cohen, J. 1976. The Global Assessment Scale: A procedure for measuring overall severity of psychiatric disturbance. Archives of general psychiatry 33(6), pp. 766–771.

21. Fagerland, M.W., Lydersen, S. and Laake, P. 2015. Recommended confidence intervals for two independent binomial proportions. Statistical Methods in Medical Research 24(2), pp. 224–254. doi: 10.1177/0962280211415469.

22. Fervaha, G., Foussias, G., Agid, O. and Remington, G. 2014. Impact of primary negative symptoms on functional outcomes in schizophrenia. European Psychiatry 29(7), pp. 449–455. doi: 10.1016/j.eurpsy.2014.01.007.

23. Frydman-Gani, C. et al. 2025. Large Language Models for Psychiatric Phenotype Extraction from Electronic Health Records. p. 2025.08.07.25333172. Available at: https://www.medrxiv.org/content/10.1101/2025.08.07.25333172v1 [Accessed: 17 September 2025].

24. Galderisi, S., Mucci, A., Buchanan, R.W. and Arango, C. 2018. Negative symptoms of schizophrenia: new developments and unanswered research questions. The Lancet Psychiatry 5(8), pp. 664–677. doi: 10.1016/S2215-0366(18)30050-6.

25. Grzenda, A. and Widge, A.S. 2024. Electronic health records and stratified psychiatry: bridge to precision treatment? Neuropsychopharmacology 49(1), pp. 285–290. doi: 10.1038/s41386-023-01724-y.

26. Hamra, G.B. 2022. Invited Commentary: Is Bias Towards the Null From Nondifferential Misclassification Wishful Thinking? American Journal of Epidemiology 191(8), pp. 1496– 1497. doi: 10.1093/aje/kwac091.

27. Harshavardhan, H., Shetty, S., Shetty, R.P., Shetty, P.U., and Sakshi. 2025. Advancements in NLP: From Rule-Based Methods to Modern Language Models. In: 2025 IEEE International Conference on Electronics, Computing and Communication Technologies (CONECCT). pp. 1–5. Available at: https://ieeexplore.ieee.org/abstract/document/11306594 [Accessed: 17 February 2026].

28. Janik, R.A. 2024. Aspects of human memory and Large Language Models. Available at: http://arxiv.org/abs/2311.03839 [Accessed: 14 April 2026].

29. Kahn, R.S. et al. 2015. Schizophrenia. Nature Reviews Disease Primers 1(1), p. 15067. doi: 10.1038/nrdp.2015.67.

30. Kestenbaum, B. 2009. Misclassification. In: Kestenbaum, B. ed. Epidemiology and Biostatistics: An Introduction to Clinical Research. New York, NY: Springer, pp. 75–89. Available at: 10.1007/978-0-387-88433-2_8 [Accessed: 9 July 2026].

31. Knevel, R. and Liao, K.P. 2023. From real-world electronic health record data to real-world results using artificial intelligence. Annals of the Rheumatic Diseases 82(3), pp. 306–311. doi: 10.1136/ard-2022-222626.

32. Koch, E. et al. 2024. How Real-World Data Can Facilitate the Development of Precision Medicine Treatment in Psychiatry. Biological Psychiatry 96(7), pp. 543–551. doi: 10.1016/j.biopsych.2024.01.001.

33. Koleck, T.A., Dreisbach, C., Bourne, P.E. and Bakken, S. 2019. Natural language processing of symptoms documented in free-text narratives of electronic health records: a systematic review. Journal of the American Medical Informatics Association 26(4), pp. 364–379. doi: 10.1093/jamia/ocy173.

34. Kreimeyer, K. et al. 2017. Natural language processing systems for capturing and standardizing unstructured clinical information: A systematic review. Journal of Biomedical Informatics 73, pp. 14–29. doi: 10.1016/j.jbi.2017.07.012.

35. Kwon, W. et al. 2023. Efficient Memory Management for Large Language Model Serving with PagedAttention. Available at: http://arxiv.org/abs/2309.06180 [Accessed: 19 December 2025].

36. Legge, S.E. et al. 2020. Clinical indicators of treatment-resistant psychosis. The British Journal of Psychiatry 216(5), pp. 259–266. doi: 10.1192/bjp.2019.120.

37. Legge, S.E. et al. 2021. Associations Between Schizophrenia Polygenic Liability, Symptom Dimensions, and Cognitive Ability in Schizophrenia. JAMA Psychiatry 78(10), pp. 1143–1151. doi: 10.1001/jamapsychiatry.2021.1961.

38. Levy, M., Jacoby, A. and Goldberg, Y. 2024. Same Task, More Tokens: the Impact of Input Length on the Reasoning Performance of Large Language Models. In: Ku, L.-W., Martins, A., and Srikumar, V. eds Proceedings of the 62nd Annual Meeting of the Association for Computational Linguistics (Volume 1: Long Papers). Bangkok, Thailand: Association for Computational Linguistics, pp. 15339–15353. Available at: https://aclanthology.org/2024.acl-long.818/ [Accessed: 14 April 2026].

39. Liu, H. and Zhang, Z. 2017. Logistic regression with misclassification in binary outcome variables: a method and software. Behaviormetrika 44(2), pp. 447–476. doi: 10.1007/s41237-017-0031-y.

40. Lynham, A.J. et al. 2023. DRAGON-Data: a platform and protocol for integrating genomic and phenotypic data across large psychiatric cohorts. BJPsych Open 9(2), pp. 1–8. doi: 10.1192/bjo.2022.636.

41. Madden, J.M., Lakoma, M.D., Rusinak, D., Lu, C.Y. and Soumerai, S.B. 2016. Missing clinical and behavioral health data in a large electronic health record (EHR) system. Journal of the American Medical Informatics Association 23(6), pp. 1143–1149. doi: 10.1093/jamia/ocw021.

42. Munzir, S.I., Hier, D.B., Oommen, C. and Carrithers, M.D. 2024. A Large Language Model Outperforms Other Computational Approaches to the High-Throughput Phenotyping of Physician Notes. AMIA … Annual Symposium proceedings. AMIA Symposium 2024, pp. 838– 846.

43. Nasrallah, H.A. 2009. Long overdue: measurement-based psychiatric practice. Current Psychiatry 8(4), pp. 14-.

44. Neuhaus, J. 1999. Bias and efficiency loss due to misclassified responses in binary regression. Biometrika 86(4), pp. 843–855. doi: 10.1093/biomet/86.4.843.

45. Pakhomov, S., Jacobsen, S.J., Chute, C.G. and Roger, V.L. 2008. Agreement between Patient-reported Symptoms and their Documentation in the Medical Record. The American journal of managed care 14(8), pp. 530–539.

46. Pogue-Geile, M.F. 1989. The Prognostic Significance of Negative Symptoms in Schizophrenia. British Journal of Psychiatry 155(S7), pp. 123–127. doi: 10.1192/S0007125000291654.

47. Quinn, T.P. et al. 2024. A primer on the use of machine learning to distil knowledge from data in biological psychiatry. Molecular Psychiatry 29(2), pp. 387–401. doi: 10.1038/s41380-023-02334-2.

48. R Core Team. 2021. R: A Language and Environment for Statistical Computing. Available at: https://www.R-project.org/.

49. Reddy, S. 2024. Generative AI in healthcare: an implementation science informed translational path on application, integration and governance. Implementation Science 19(1), p. 27. doi: 10.1186/s13012-024-01357-9.

50. Salvatore, N., Wang, H. and Zhang, Q. 2025. Lost in the Middle: An Emergent Property from Information Retrieval Demands in LLMs. Available at: http://arxiv.org/abs/2510.10276 [Accessed: 14 April 2026].

51. Smoller, J.W. 2018. The use of electronic health records for psychiatric phenotyping and genomics. American Journal of Medical Genetics Part B: Neuropsychiatric Genetics 177(7), pp. 601–612. doi: 10.1002/ajmg.b.32548.

52. Tian, S. et al. 2024. Opportunities and challenges for ChatGPT and large language models in biomedicine and health. Briefings in Bioinformatics 25(1), p. bbad493. doi: 10.1093/bib/bbad493.

53. Van Rossum, G. and Drake, F.L. 2009. Python 3 Reference Manual. Scotts Valley, CA: CreateSpace.

54. Varshney, N., Raj, S., Mishra, V., Chatterjee, A., Saeidi, A., Sarkar, R. and Baral, C. 2025. Investigating and Addressing Hallucinations of LLMs in Tasks Involving Negation. In: Cao, T. et al. eds Proceedings of the 5th Workshop on Trustworthy NLP (TrustNLP 2025). Albuquerque, New Mexico: Association for Computational Linguistics, pp. 580–598. Available at: https://aclanthology.org/2025.trustnlp-main.37/ [Accessed: 10 April 2026].

55. Wing, J.K. et al. 1990. SCAN: Schedules for Clinical Assessment in Neuropsychiatry. Archives of General Psychiatry 47(6), pp. 589–593. doi: 10.1001/archpsyc.1990.01810180089012.

56. Wright, A.G.C., Ringwald, W.R., Vize, C.E., Eichstaedt, J.C., Angstadt, M., Taxali, A. and Sripada, C. 2026. Assessing personality using zero-shot generative AI scoring of brief open-ended text. Nature Human Behaviour, pp. 1–15. doi: 10.1038/s41562-025-02389-x.

