## Supplementary Materials for "Extracting Symptoms of Psychotic Disorders from Clinical Notes using Natural Language Processing"

### SUPPLEMENTARY METHODS

#### Symptom Extraction using Encoder Models

##### *Data Preprocessing*

Case note summaries and gold standard labels were loaded using the Hugging Face datasets library. Case note summaries were fed to a model specific AutoTokenizer, converting text data to numeric input IDs and creating an attention mask. Gold standard labels were aligned, and missing values were mapped to -100 to ensure that they are ignored during training. The output for each record was a dictionary containing input IDs, attention masks, and aligned gold-standard labels, which were then batched for training.

##### *Data Splitting Strategy*

Case notes were summarised by over 17 different individuals to form Case note summaries. Some case notes were summarised by more than one summariser; summariser groups were formed from these. To avoid data leakage, the dataset was split at the “summariser group” level rather than randomly. Scheme 1 (S1) data was kept separate from non-S1 data throughout the split. Thus, a patient record from the training set never appeared in the validation or test sets. The validation and test sets are split in roughly a 2:1 ratio.

##### *Classification*

We used Pre-trained Language Models (PLMs), including standard models like BERT and RoBERTa, as well as clinical domain-specific variants, to make predictions from clinical notes.

We trained one independent binary classifier per clinical symptom per model. The input text was tokenised and passed through the model encoder, which produced a context-aware representation of the whole document (taken from the [CLS] token). This representation passed through a dropout layer and a linear classification head to produce a set of raw output scores (logits), one per label. Optuna (Akiba et al. 2019) was used for automated hyperparameter tuning, running 30 trials to find the best combination of learning rate, batch size, and gradient accumulation steps.

Formally, given an input text sequence $(X = x_1, x_2, \dots, x_n)$, the sequence was tokenized and passed through the PLM encoder to yield contextualized hidden states. We extracted the aggregate document representation $h_{pooled} \in R^d$ (typically derived from the $[CLS]$ token, where $d$ is the hidden dimension size).

To generate predictions for $L$ distinct clinical labels, $h_{pooled}$ was passed through a dropout layer and a randomly initialized linear classification head:

$z = W \cdot Dropout(h_{pooled}) + b$

where $W \in R^{L \times d}$ and $b \in R^{L}$ are the trainable weight matrix and bias, and $z$ represents the raw output logits for all labels.

##### *Handling Long Clinical Documents*

Clinical notes are often longer than the 512-token limit that standard models like BERT can handle. Mean-Pooling Chunking was used to handle this. The document was split into up to 8 sequential chunks, each within 512 tokens. Each chunk was processed independently by the model to produce its own logit. These chunk-level logits were then averaged to give a single document-level prediction.

Formally, the document was segmented into $K$ sequential chunks (e.g., $K$ = 8), each up to 512 tokens long. Each chunk is independently processed by the PLM to produce a chunk-level logit $z_K$. The final document-level logit $z_{final}$ is obtained via mean-pooling:

$$z_{final} = \frac{1}{K} \sum_{k=1}^{K} z_k$$

##### *Addressing Extreme Class Imbalance*

Where there is class imbalance, a standard cross-entropy loss would push the model to predict the most frequent class (i.e., where ‘absent’ ratings are rare for delusions, the model would always predict ‘present’). To fix this, we replaced the standard loss with a customised Focal Loss.

##### *Model Inference*

During the evaluation phase, the pipeline dynamically loads the corresponding fine-tuned binary model for each specific label. The model processes the unseen test data to output raw logits ($z$).

To generate the final prediction, the logit was passed through a Sigmoid activation function to map it to a probability score between 0 and 1:

$P(y = 1 \mid x) = \frac{1}{1 + e^{-z}}$

Rather than relying on a default 0.5 cut off, we used an optimal threshold (τ) that was dynamically calculated during the validation phase (optimized specifically for the F1-score). The final hard prediction (0 or 1) is made by comparing the probability against this optimal threshold:

$$y_{\text{pred}} = \begin{cases} 1, & \text{if } P(y = 1 \mid x) > \tau \\ 0, & \text{otherwise} \end{cases}$$

##### *Model Evaluation*

The pipeline was run under two complementary experimental setups to ensure reliable results and coverage across all data points.

In the first setup, all records from the S1 summarise group were used as the training set. Records from the remaining summarise groups were then split into a validation set and a test set. The model was trained exclusively on S1 data and evaluated on the held-out non-S1 data. In the second setup, the roles were reversed. Records from all non-S1 summarise groups were used as the training set, and the S1 summarise group was used as the validation and test set.

The final reported results combine the test outputs from both setups. Because each setup uses a different portion of the data as the test set, together they cover all records in the dataset. This means every data point has been evaluated exactly once.

### SUPPLEMENTARY FIGURES

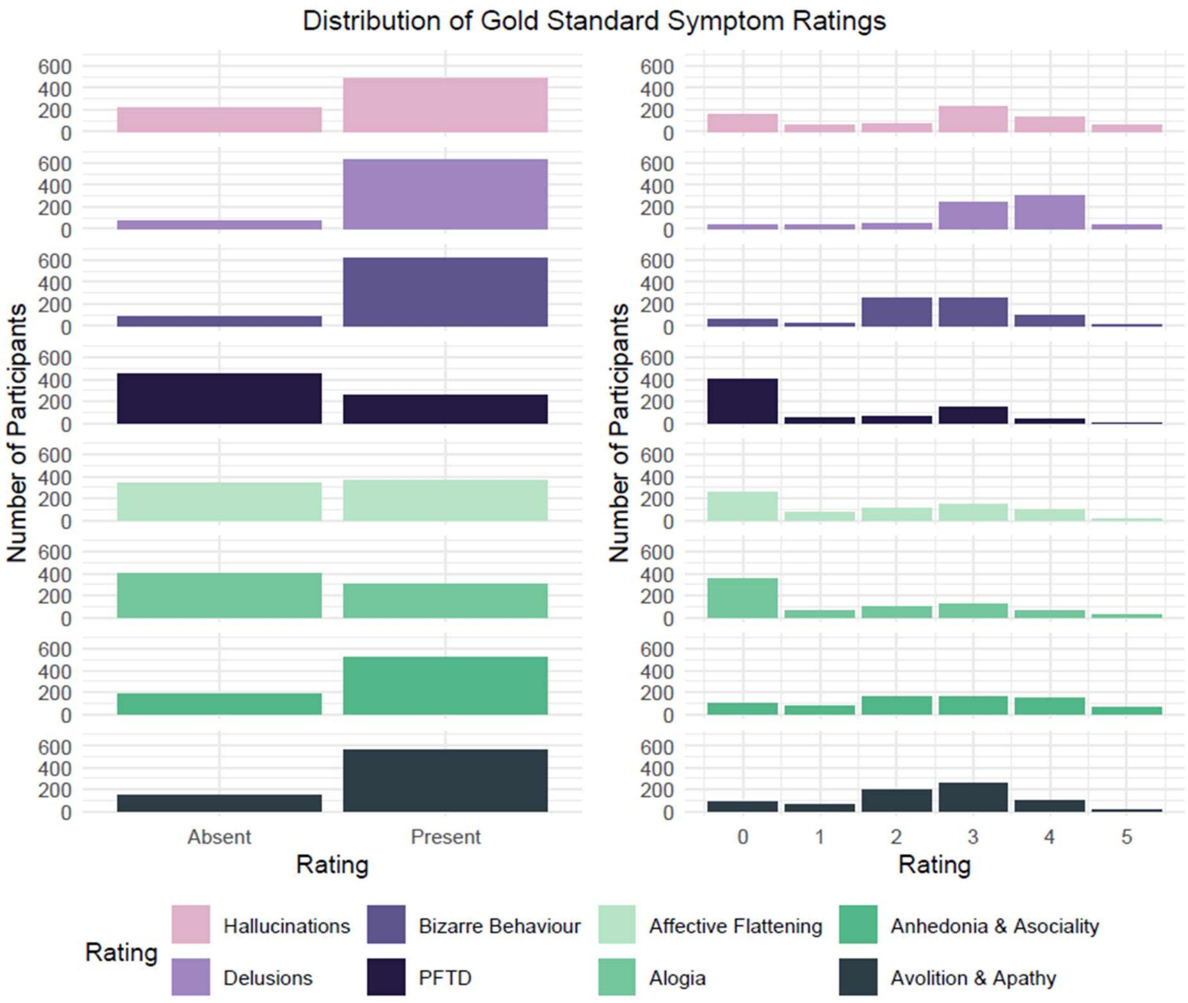

Supplementary Figure 1. Distribution of gold-standard phenotypes. Plots on the left represent frequency of each symptom's presence and absence in the gold-standard ratings. Plots on the right represent the frequencies of each symptom's ratings on the original SAPS and SANS scale, where scores of 0 represent symptom absence and scores of 5 represent severe manifestations of that symptom.

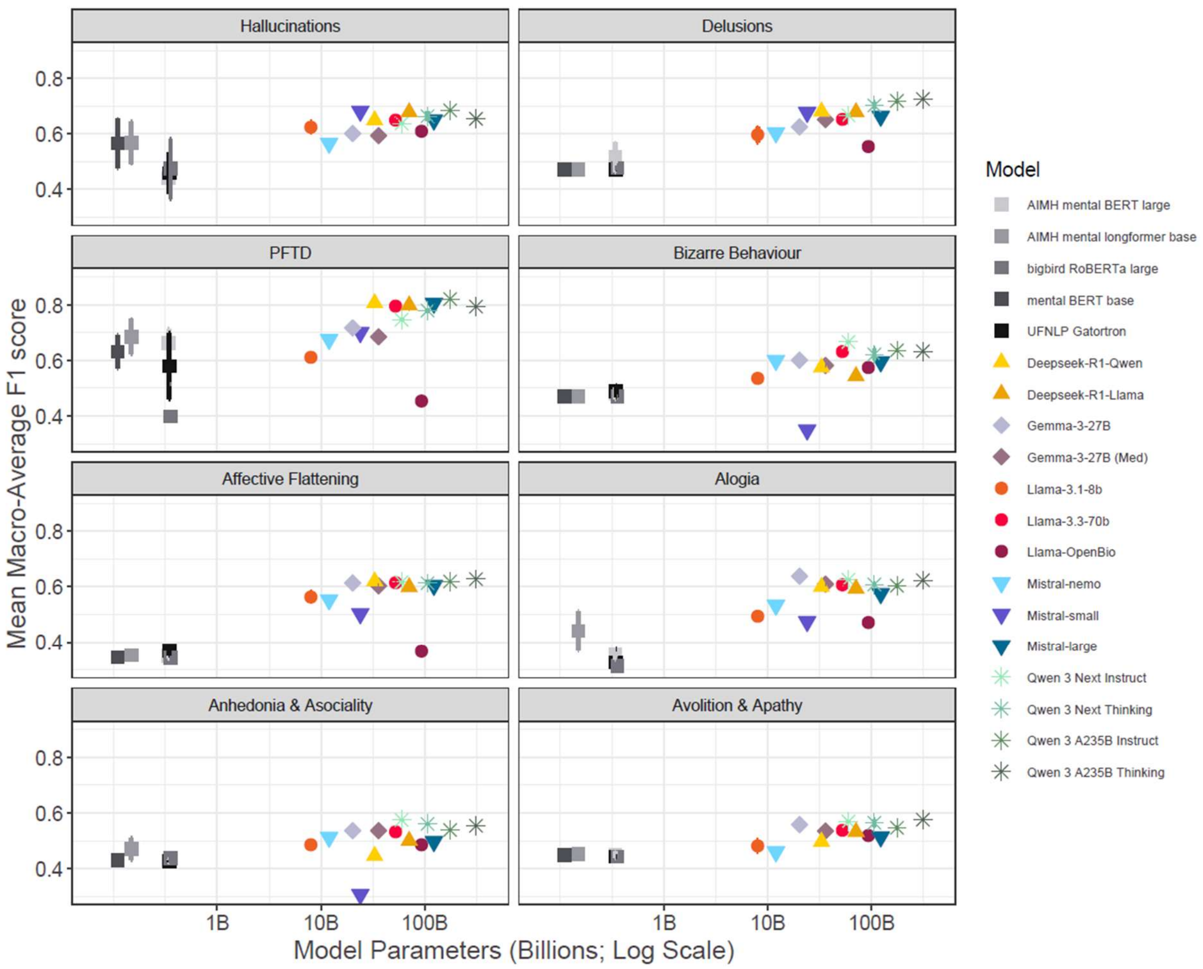

Supplementary Figure 2. Full comparison of Mean Macro-Average F1 Score across different symptoms and models in a zero-shot setting. Model size, indexed by number of parameters, is represented on the log scale on the x-axis. Error bars represent standard deviation around the mean.  Each model is given a different colour, and each model family given a different shape.

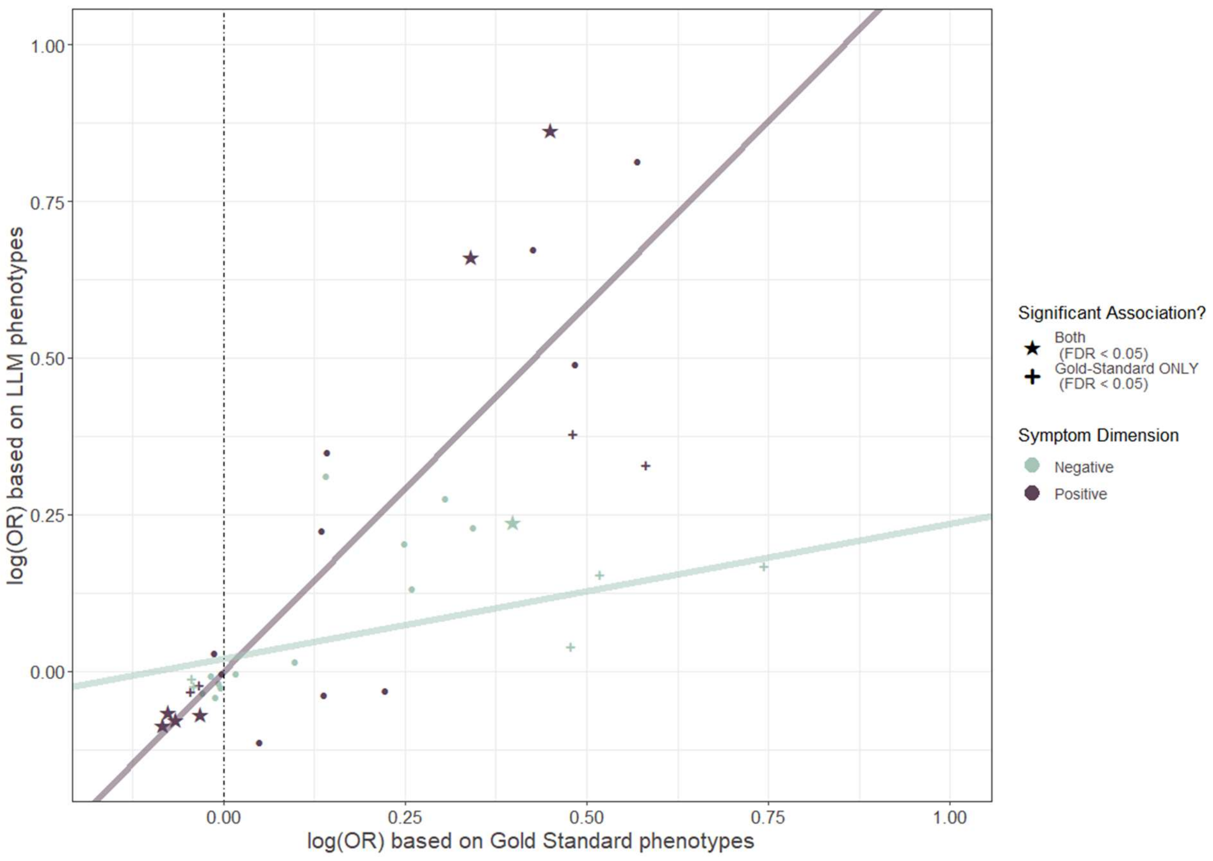

Supplementary Figure 3. Relationship between log(OR) for associations using LLM-predicted vs gold-standard phenotypes. Effect sizes for associations with gold-standard phenotypes are displayed across the x-axis, whereas those with LLM-predicted phenotypes are across the y-axis. Stars represent associations that are significant (FDR < 0.05) in both analyses, whereas crosses represent associations that are significant only in the gold-standard analysis. All other associations are represented by points. Data are coloured by symptom dimension, where positive symptoms (i.e., hallucinations, delusions, PFTD, bizarre behaviour) are deep purple, and negative symptoms (i.e., affective flattening, alogia, anhedonia and asociality, avolition and apathy) are light green. Lines represent the slope of associations between the gold-standard and LLM-predicted effect sizes for positive and negative symptom dimensions, separately.

### SUPPLEMENTARY TABLES

Symptom definitions used in the LLM analysis.

| Symptom | Definition | SAPS/SANS item |
|---|---|---|
| Hallucinations | HALLUCINATIONS refer to perceptual experiences occurring without external stimuli. Ensure that ONLY false perceptions are rated as hallucinations, not false thoughts or beliefs.<br><br>INCLUDE the following when documented in clinical notes:<br>- Auditory hallucinations (AH): Patient reports hearing voices, sounds, or noises not heard by others. Common presentations include: voices commenting on patient's actions or thoughts, two or more voices conversing, command hallucinations or voices giving instructions, and documentation phrases such as "patient reports hearing voices", "AH present", "responding to internal stimuli", "observed talking to self when alone".<br>- Visual hallucinations (VH): Patient reports seeing figures, shapes, shadows, or people not visible to others<br>- Olfactory hallucinations: Patient reports smelling odours others cannot detect, or believes they emit odours despite reassurance<br>- Gustatory hallucinations: Patient reports unusual tastes without corresponding stimuli<br>- Tactile/somatic hallucinations: Patient reports physical sensations (crawling, burning, being touched) without cause<br><br>EXCLUDE the following:<br>- Pseudohallucinations (patient recognises experiences as internally generated, often documented as "patient acknowledges voices are not real")<br>- Illusions (misperceptions of real stimuli, e.g., mistaking a coat for a person)<br>- Hypnagogic/hypnopompic experiences (hallucinations when falling asleep or waking)<br>- Flashbacks explicitly attributed to PTSD/trauma<br>- Hallucinations explicitly attributed to substance intoxication/withdrawal in that specific episode<br>- Dreams or nightmares<br>- Intrusive thoughts (unless experienced as external voices) | SAPS Global Hallucinations (Lifetime) |

| | | |
|---|---|---|
| Delusions | DELUSIONS are fixed, false beliefs maintained despite contradictory evidence. These beliefs should be firmly held, fixed, and complex.<br><br>INCLUDE the following when documented in clinical notes:<br>- Persecutory/paranoid delusions: Belief of being harassed, followed, poisoned, spied upon, or conspired against<br> - Documentation phrases: "paranoid ideation", "believes neighbours are monitoring him", "persecutory beliefs"<br>- Grandiose delusions: Inflated sense of power, knowledge, identity, or special relationship to deity/famous person<br>- Delusions of reference: Belief that events, objects, or people have particular personal significance (e.g., TV sending messages)<br>- Delusions of control/passivity: Belief that thoughts, feelings, or actions are controlled by external force<br>- Thought broadcasting: Belief that thoughts can be heard by others<br>- Thought insertion: Belief that thoughts are placed in one's mind by external source<br>- Thought withdrawal: Belief that thoughts are being removed<br>- Somatic delusions: False beliefs about body (e.g., organs rotting, infested with parasites) NOT explained by medical condition<br>- Delusions of jealousy: Unfounded conviction of partner's infidelity<br>- Nihilistic delusions: Belief that self, others, or world does not exist or is ending<br>- Religious delusions: Delusions with religious content beyond cultural norms (e.g., being a prophet, special mission from God)<br><br>EXCLUDE the following:<br>- Overvalued ideas (strongly held beliefs that are amenable to reason)<br>- Suspiciousness that is proportionate to circumstances (e.g., genuine persecution, realistic concerns)<br>- Culturally normative beliefs (consider patient's religious/cultural context)<br>- Beliefs explicitly documented as "insight intact" or "patient recognises beliefs were delusional"<br>- Confabulation in context of documented memory impairment | SAPS Global Delusions (Lifetime) |

| | | |
|---|---|---|
| | - Misidentification due to dementia or delirium<br>- Fleeting thoughts or paranoia for example, the belief that everyone is talking about them without knowing who or why.<br>- False perceptions | |
| Positive Formal Thought Disorder | POSITIVE FORMAL THOUGHT DISORDER refers to disorganisation in the form (not content) of thinking, as manifested in speech and documented by clinical observations or direct quotes. For a rating of positive formal thought disorder, at least one of the following symptoms must be present: derailment, tangentiality, incoherence, illogicality, circumstantiality, or pressure of speech. A mention of disorientation alone is not sufficient to rate positive formal thought disorder as present. Additionally, lack of insight into one's disorder is not evidence of positive formal thought disorder.<br><br>INCLUDE the following when documented in clinical notes:<br>- Derailment/loose associations: Ideas slip from one track to another with little or no connection<br> - Documentation phrases: "loosening of associations" "LOA" "tangential speech" "thought disordered"<br>- Tangentiality: Replies to questions that are oblique or irrelevant<br>- Incoherence/word salad: Speech that is incomprehensible due to lack of logical connection<br> - Documentation phrases: "incoherent speech" "word salad" "incomprehensible"<br>- Illogicality: Conclusions reached that do not follow logically<br>- Circumstantiality: Speech that is delayed in reaching the point due to excessive unnecessary detail (but eventually reaches it)<br>- Pressure of speech: Rapid, excessive speech that is difficult to interrupt<br> - Documentation phrases: "pressured speech"<br>- Clanging: Speech guided by sound rather than meaning<br>- Neologisms: Made-up words or phrases<br><br>EXCLUDE the following:<br>- Disorganised speech due to aphasia, dysarthria, or other neurological conditions<br>- Speech abnormalities attributed to intellectual disability<br>- Disorganisation explicitly attributed to intoxication or delirium | SAPS Global Positive Thought Disorder (Lifetime) |

| | | |
|---|---|---|
| | - Circumstantiality alone without other features (common in anxiety, mania) or just the term "flight of ideas"<br>- Non-native English speaker with language difficulties<br>- Normal disfluencies or difficulty finding words<br>- Descriptions of delusions (e.g., thought insertion and thought withdrawal) and bizarre behaviour | |
| Bizarre behaviour | BIZARRE BEHAVIOUR refers to clearly unusual, bizarre or odd behaviour.<br><br>INCLUDE the following when documented in clinical notes:<br>- Unusual appearance/dress: Bizarre clothing choices, inappropriate dress for weather/context, unusual makeup or adornments<br>- Odd social or sexual behaviour: Socially inappropriate actions, violation of social norms not due to ignorance<br>- Stereotyped behaviour: Repetitive, non-functional movements or rituals<br>- Aggressive/agitated behaviour: When bizarre in nature or grossly disproportionate to circumstances or unprovoked<br>- Odd ritualistic behaviour: Unusual routines or behaviours with no clear purpose<br>- Responding to hallucinations<br><br>EXCLUDE the following:<br>- Behaviour directly attributable to substance intoxication or withdrawal<br>- Agitation or aggression with clear precipitant and proportionate response<br>- Poor self-care due to depression or negative symptoms (rate under avolition/apathy)<br>- The term "bizarre" in the context of other types of symptoms (e.g., bizarre delusions) | SAPS Global Bizarre Behaviour (Lifetime) |
| Affective Flattening | AFFECTIVE FLATTENING (also termed blunted affect or flat affect) refers to a reduction in the range and intensity of emotional expression. Affective flattening relates to reduced emotional expression not reduced emotional experience (i.e., flat mood).<br><br>INCLUDE the following when documented in clinical notes:<br>- Reduced facial expressivity: Unchanging facial expression, reduced emotional display<br> - Documentation phrases: "flat affect" "blunted affect" "restricted affect" "reduced affective range" | SANS Global Affective Flattening (Lifetime) |

| | | |
|---|---|---|
|  | - Decreased spontaneous movement: Sits motionless, reduced gestures during conversation<br>- Poor eye contact: Avoids or fails to make eye contact (when not culturally normative)<br>- Affective non-responsivity: Fails to respond emotionally to emotionally evocative material<br>- Inappropriate affect: Emotional display incongruent with content (e.g., laughing when discussing sad events)<br> - Documentation phrases: "inappropriate affect" "incongruent affect"<br>- Reduced vocal inflection: Monotone speech, lack of prosody<br> - Documentation phrases: "monotonous speech" "lacks vocal inflection"<br><br>EXCLUDE the following:<br>- Depressed affect (sadness, tearfulness) - this is restricted but not flat<br>- Affect abnormalities explicitly attributed to medication side effects (e.g., antipsychotic-induced)<br>- Affect restricted to clinical interview in context of suspiciousness/paranoia |  |
| Alogia | ALOGIA refers to a reduction in the quantity or content of speech.<br>INCLUDE the following when documented in clinical notes:<br>- Poverty of speech: Reduced amount of spontaneous speech, brief/unelaborated responses<br>- Poverty of content: Speech is adequate in amount but conveys little information, vague, overly abstract or concrete<br>- Increased latency of response: Long delays before responding to questions<br>- Blocking: Sudden interruption in train of thought, patient cannot recall what they were saying<br><br>EXCLUDE the following:<br>- Mutism (Alogia is a reduction in the amount of speech rather than a complete loss of speech).<br>- Lack of vocal inflection or tone | SANS Global Alogia (Lifetime) |
| Anhedonia and Asociality | ANHEDONIA AND ASOCIALITY refer to reduced ability to experience pleasure and reduced social drive. This is seen as a lack of interest in relationships.<br><br>INCLUDE the following when documented in clinical notes: | SANS Global Anhedonia and Asociality (Lifetime) |

| | | |
|---|---|---|
| | - Reduced social relationships: Few or no friends, does not seek social contact, socially isolated<br>- Reduced interest in relationships: Does not desire friendships or intimate relationships<br>- Inability to feel intimacy/closeness: Feels distant from family and friends despite contact<br>- Reduced recreational interests: Previously enjoyed activities no longer provide pleasure<br>- Reduced sexual interest: Decreased libido or interest in sexual activity (consider age-appropriateness)<br>- Social withdrawal: Spends most time alone, avoids social situations<br><br>EXCLUDE the following:<br>- Problems with appearance and hygiene or bizarre behaviour | |
| Avolition and Apathy | AVOLITION AND APATHY refers to reduced motivation and drive to initiate and persist in goal-directed activities.<br><br>INCLUDE the following when documented in clinical notes:<br>- Physical anergia: Lack of energy, tends to sit for long periods, sleeps excessively during day<br>- Poor grooming and hygiene: Does not bathe, change clothes, or attend to personal care<br>- Impersistence at work/school: Cannot maintain employment or educational activities, drops out<br><br>EXCLUDE the following:<br>- Anergia explicitly attributed to depression | SANS Global Avolition and Apathy (Lifetime) |

Supplementary Table 1. Symptom definitions used in zero-shot and few-shot prompts for LLMs.

#### Zero- and few-shot prompts used in the LLM analysis.

| Zero-Shot | Few-Shot |
|---|---|
| You are a world-leading psychiatrist with expertise in schizophrenia and symptoms of psychosis. Use the patient note to answer the question. Respond briefly with ONLY "1" or "0".<br><br>The following is a psychiatric patient note for a symptom classification task. Please scan the text for keywords related to {symp_name} and identify any descriptions matching the definition. {definition}. Based on your analysis, has this patient shown evidence of {symp_name} at any point throughout the patient note? Respond "1" if there is evidence that the patient has experienced {symp_name}, even if it appears in an historical context or is described synonymously (i.e., not explicitly stated by name). Respond "0" if there is no evidence that the patient experienced {symp_name}. Thank you.<br><br>Patient note: {note} | You are a world-leading psychiatrist with expertise in schizophrenia and symptoms of psychosis. Use the patient note to answer the question. Respond briefly with ONLY "1" or "0".<br><br>The following is a psychiatric patient note for a symptom classification task. Please scan the text for keywords related to {symp_name} and identify any descriptions matching the definition. {definition} Use the following examples to help. Example 1: {e1} // Rating 1: {r1}. Example 2: {e2} // Rating 2: {r2}. Example 3: {e3} // Rating 3: {r3}. Example 4: {e4} // Rating 4: {r4}. Based on your analysis, is there evidence of {symp_name} at any point throughout the patient note? Respond "1" if there is evidence of {symp_name}, even if it appears in an historical context or is described synonymously (i.e., not explicitly stated by name). Respond "0" if there is no evidence of {symp_name}. Thank you."""<br><br>Patient note: {note} |

Supplementary Table 2. Prompts used for zero-shot and few-shot analyses. {symp_name} reflects one of the eight symptoms outlined; {definition} for each symptom can be found **in Supplementary Table 1** and is derived from the SAPS and SANS ratings guides used during CardiffCOGS data collection (Andreasen 1984; Andreasen 1989). {note} reflects a clinical summary from an individual in the CardiffCOGS sample. In the few-shot example {e1} reflects a short extract from a clinical summary and {r1} reflects the gold-standard that would be given for it.

Definitions of Evaluation Metrics.

| Metric | Equation | Definition |
|---|---|---|
| Precision | $\frac{TP}{TP + FP}$ | The ratio of true positives to the total number of model-predicted positives. |
| Sensitivity ("Recall") | $\frac{TP}{TP + FN}$ | The ratio of true positives to the total number of all positives. This may also be called "Recall" or "True Positive Rate". |
| Specificity | $\frac{TN}{TN + FP}$ | Ratio of true negatives to the total number of all negatives. |
| F1 | $\frac{2 \times TP}{2 \times TP + FP + FN}$ or $\frac{2 \times precision \times recall}{precision + recall}$ | Harmonic mean of precision and recall. |
| Accuracy | $\frac{TP + TN}{TP + TN + FP + FN}$ | Proportion of correctly predicted examples. |
| Balanced Accuracy (BAC) | $\frac{Sensitivity + Specificity}{2}$ | A measure of accuracy that is more robust to class imbalance. |
| Macro-Average F1 | $\frac{\sum F1i}{n}$ | Average F1 across all classes (i.e., for binary predictions the average of $F1_{present}$ and $F1_{absent}$). |
| Matthew's Correlation Coefficient (MCC) | $\frac{TP \times TN - FP \times FN}{\sqrt{(TP + FP) \times (TP + FN) \times (TN + FP) \times (TN + FN)}}$ | MCC of 1 indicates perfect agreement between predictions and gold standard. MCC = -1 indicates absolute disagreement. MCC = 0 = chance. |

Supplementary Table 3. Definitions of evaluation metrics used to assess LLM performance. Table adapted from Quinn et al. (2024). TP = True Positives; TN = True Negatives; FP = False Positives; FN = False Negatives; MCC = Matthews Correlation Coefficient.

#### Tables of results for Pretrained Language Models (PLMs)

| Model | Precision | Recall/Sensitivity | Specificity | F1 (present) | Macro-Average | Accuracy | BAC | MCC |
|---|---|---|---|---|---|---|---|---|
| AIMH_mental-bert-large-uncased-binary | 0.689 | 0.965 | 0.096 | 0.794 | 0.460 | 0.706 | 0.530 | 0.084 |
| AIMH_mental-longformer-base-4096-binary | 0.702 | 0.934 | 0.151 | 0.793 | 0.489 | 0.717 | 0.543 | 0.103 |
| UFNLP_gatortron-base-binary | 0.684 | 0.960 | 0.081 | 0.787 | 0.445 | 0.695 | 0.520 | 0.051 |
| google_bigbird-roberta-large-binary | 0.672 | 0.988 | 0.035 | 0.785 | 0.419 | 0.678 | 0.512 | 0.038 |
| mental_mental-bert-base-uncased-binary | 0.690 | 0.963 | 0.099 | 0.793 | 0.458 | 0.704 | 0.531 | 0.079 |

Supplementary Table 4. Comparing Mean Metrics across symptoms for Pretrained Language Models.

| Model | Precision | Recall/Sensitivity | Specificity | F1 (present) | Macro-Average | Accuracy | BAC | MCC |
|---|---|---|---|---|---|---|---|---|
| AIMH_mental-bert-large-uncased | 0.701 | 0.986 | 0.033 | 0.819 | 0.440 | 0.697 | 0.510 | 0.053 |
| AIMH_mental-longformer-base-4096 | 0.739 | 0.919 | 0.247 | 0.817 | 0.569 | 0.715 | 0.583 | 0.235 |
| UFNLP_gatortron-base | 0.705 | 0.968 | 0.066 | 0.815 | 0.459 | 0.694 | 0.517 | 0.049 |
| google_bigbird-roberta-large | 0.713 | 0.970 | 0.095 | 0.821 | 0.475 | 0.705 | 0.533 | 0.102 |
| mental_mental-bert-base-uncased | 0.736 | 0.953 | 0.210 | 0.830 | 0.566 | 0.727 | 0.581 | 0.244 |

Supplementary Table 5. Full Pretrained Language Model results for detecting presence or absence of Hallucinations.

| Model | Precision | Recall/Sensitivity | Specificity | F1 (present) | Macro-Average | Accuracy | BAC | MCC |
|---|---|---|---|---|---|---|---|---|
| AIMH_mental-bert-large-uncased | 0.901 | 0.995 | 0.050 | 0.946 | 0.515 | 0.897 | 0.523 | 0.109 |
| AIMH_mental-longformer-base-4096 | 0.896 | 1.000 | 0.000 | 0.945 | 0.473 | 0.896 | 0.500 | 0.000 |
| UFNLP_gatortron-base | 0.896 | 1.000 | 0.000 | 0.945 | 0.473 | 0.896 | 0.500 | 0.000 |
| google_bigbird-roberta-large | 0.897 | 0.999 | 0.005 | 0.945 | 0.477 | 0.896 | 0.502 | 0.023 |
| mental_mental-bert-base-uncased | 0.896 | 1.000 | 0.000 | 0.945 | 0.473 | 0.896 | 0.500 | 0.000 |

Supplementary Table 6. Full Pretrained Language Model results for detecting presence or absence of Delusions.

| Model | Precision | Recall/Sensitivity | Specificity | F1 (present) | Macro-Average | Accuracy | BAC | MCC |
|---|---|---|---|---|---|---|---|---|
| AIMH_mental-bert-large-uncased | 0.537 | 0.804 | 0.585 | 0.640 | 0.662 | 0.665 | 0.694 | 0.382 |
| AIMH_mental-longformer-base-4096 | 0.579 | 0.698 | 0.695 | 0.630 | 0.685 | 0.696 | 0.696 | 0.383 |
| UFNLP_gatortron-base | 0.493 | 0.765 | 0.497 | 0.584 | 0.578 | 0.595 | 0.631 | 0.278 |
| google_bigbird-roberta-large | 0.396 | 0.943 | 0.156 | 0.557 | 0.398 | 0.446 | 0.549 | 0.127 |
| mental_mental-bert-base-uncased | 0.515 | 0.752 | 0.572 | 0.605 | 0.631 | 0.638 | 0.662 | 0.321 |

Supplementary Table 7. Full Pretrained Language Model results for detecting presence or absence of Positive Formal Thought Disorder.

| Model | Precision | Recall/Sensitivity | Specificity | F1 (present) | Macro-Average | Accuracy | BAC | MCC |
|---|---|---|---|---|---|---|---|---|
| AIMH_mental-bert-large-uncased | 0.888 | 0.999 | 0.013 | 0.940 | 0.482 | 0.887 | 0.506 | 0.051 |
| AIMH_mental-longformer-base-4096 | 0.886 | 1.000 | 0.000 | 0.940 | 0.470 | 0.886 | 0.500 | 0.000 |
| UFNLP_gatortron-base | 0.888 | 0.995 | 0.021 | 0.939 | 0.488 | 0.884 | 0.508 | 0.063 |
| google_bigbird-roberta-large | 0.886 | 1.000 | 0.000 | 0.940 | 0.470 | 0.886 | 0.500 | 0.000 |
| mental_mental-bert-base-uncased | 0.886 | 1.000 | 0.000 | 0.940 | 0.470 | 0.886 | 0.500 | 0.000 |

Supplementary Table 8. Full Pretrained Language Model results for detecting presence or absence of Bizarre Behaviour.

| Model | Precision | Recall/Sensitivity | Specificity | F1 (present) | Macro-Average | Accuracy | BAC | MCC |
|---|---|---|---|---|---|---|---|---|
| AIMH_mental-bert-large-uncased | 0.522 | 0.992 | 0.009 | 0.684 | 0.350 | 0.521 | 0.500 | 0.003 |
| AIMH_mental-longformer-base-4096 | 0.523 | 0.995 | 0.010 | 0.685 | 0.352 | 0.523 | 0.502 | 0.015 |
| UFNLP_gatortron-base | 0.522 | 0.970 | 0.031 | 0.678 | 0.368 | 0.520 | 0.500 | 0.000 |
| google_bigbird-roberta-large | 0.521 | 1.000 | 0.000 | 0.685 | 0.343 | 0.521 | 0.500 | 0.000 |
| mental_mental-bert-base-uncased | 0.522 | 1.000 | 0.002 | 0.686 | 0.345 | 0.522 | 0.501 | 0.026 |

Supplementary Table 9. Full Pretrained Language Model results for detecting presence or absence of Affective Flattening.

| Model | Precision | Recall/Sensitivity | Specificity | F1 (present) | Macro-Average | Accuracy | BAC | MCC |
|---|---|---|---|---|---|---|---|---|
| AIMH_mental-bert-large-uncased | 0.428 | 0.952 | 0.066 | 0.591 | 0.354 | 0.441 | 0.509 | 0.047 |
| AIMH_mental-longformer-base-4096 | 0.449 | 0.900 | 0.184 | 0.599 | 0.441 | 0.488 | 0.542 | 0.113 |
| UFNLP_gatortron-base | 0.428 | 0.984 | 0.031 | 0.596 | 0.326 | 0.435 | 0.508 | 0.029 |
| google_bigbird-roberta-large | 0.426 | 0.996 | 0.015 | 0.597 | 0.313 | 0.431 | 0.505 | 0.029 |
| mental_mental-bert-base-uncased | 0.424 | 1.000 | 0.000 | 0.595 | 0.298 | 0.424 | 0.500 | 0.000 |

Supplementary Table 10. Full Pretrained Language Model results for detecting presence or absence of Alogia.

| Model | Precision | Recall/Sensitivity | Specificity | F1 (present) | Macro-Average | Accuracy | BAC | MCC |
|---|---|---|---|---|---|---|---|---|
| AIMH_mental-bert-large-uncased | 0.745 | 1.000 | 0.002 | 0.854 | 0.429 | 0.745 | 0.501 | 0.021 |
| AIMH_mental-longformer-base-4096 | 0.750 | 0.967 | 0.061 | 0.845 | 0.472 | 0.736 | 0.514 | 0.054 |
| UFNLP_gatortron-base | 0.744 | 0.999 | 0.000 | 0.853 | 0.427 | 0.744 | 0.499 | -0.015 |
| google_bigbird-roberta-large | 0.746 | 0.996 | 0.011 | 0.853 | 0.437 | 0.744 | 0.503 | 0.022 |
| mental_mental-bert-base-uncased | 0.745 | 0.998 | 0.004 | 0.853 | 0.430 | 0.744 | 0.501 | 0.009 |

Supplementary Table 11. Full Pretrained Language Model results for detecting presence or absence of Anhedonia & Asociality.

| Model | Precision | Recall/Sensitivity | Specificity | F1 (present) | Macro-Average | Accuracy | BAC | MCC |
|---|---|---|---|---|---|---|---|---|
| AIMH_mental-bert-large-uncased | 0.794 | 0.992 | 0.009 | 0.882 | 0.449 | 0.790 | 0.501 | 0.004 |
| AIMH_mental-longformer-base-4096 | 0.795 | 0.998 | 0.009 | 0.885 | 0.451 | 0.794 | 0.503 | 0.026 |
| UFNLP_gatortron-base | 0.794 | 1.000 | 0.000 | 0.885 | 0.443 | 0.794 | 0.500 | 0.000 |
| google_bigbird-roberta-large | 0.794 | 1.000 | 0.000 | 0.885 | 0.443 | 0.794 | 0.500 | 0.000 |
| mental_mental-bert-base-uncased | 0.795 | 0.999 | 0.007 | 0.886 | 0.450 | 0.795 | 0.503 | 0.034 |

Supplementary Table 12. Full Pretrained Language Model results for detecting presence or absence of Avolition & Apathy.

#### Tables of results for zero-shot prompts and Large Language Models (LLMs)

| Model | Precision | Recall/Sensitivity | Specificity | F1 (present) | Macro-Average | Accuracy | BAC | MCC |
|---|---|---|---|---|---|---|---|---|
| Deepseek-R1-Llama | 0.809 | 0.616 | 0.698 | 0.688 | 0.615 | 0.678 | 0.657 | 0.300 |
| Deepseek-R1-Qwen | 0.808 | 0.615 | 0.693 | 0.680 | 0.609 | 0.673 | 0.654 | 0.298 |
| Gemma-3-27B | 0.765 | 0.725 | 0.550 | 0.730 | 0.611 | 0.693 | 0.638 | 0.272 |
| Gemma-3-27B (Med) | 0.744 | 0.736 | 0.517 | 0.727 | 0.599 | 0.677 | 0.626 | 0.250 |
| Llama-3.1-8b | 0.712 | 0.666 | 0.484 | 0.674 | 0.549 | 0.619 | 0.575 | 0.140 |
| Llama-3.3-70b | 0.778 | 0.712 | 0.574 | 0.734 | 0.627 | 0.710 | 0.643 | 0.291 |
| Llama-OpenBio | 0.685 | 0.952 | 0.144 | 0.784 | 0.504 | 0.690 | 0.548 | 0.153 |
| Mistral-large | 0.792 | 0.646 | 0.652 | 0.698 | 0.613 | 0.684 | 0.649 | 0.288 |
| Mistral-nemo | 0.726 | 0.671 | 0.508 | 0.680 | 0.563 | 0.648 | 0.590 | 0.182 |
| Mistral-small | 0.856 | 0.371 | 0.858 | 0.453 | 0.493 | 0.558 | 0.615 | 0.245 |
| Qwen 3 A235B Instruct | 0.808 | 0.685 | 0.648 | 0.733 | 0.646 | 0.719 | 0.667 | 0.331 |
| Qwen 3 A235B Thinking | 0.790 | 0.716 | 0.628 | 0.745 | 0.648 | 0.718 | 0.672 | 0.325 |
| Qwen 3 Next Instruct | 0.770 | 0.749 | 0.553 | 0.751 | 0.638 | 0.719 | 0.651 | 0.303 |
| Qwen 3 Next Thinking | 0.776 | 0.729 | 0.593 | 0.746 | 0.639 | 0.713 | 0.661 | 0.302 |

Supplementary Table 13. Comparing Mean Metrics across symptoms for Large Language Models (Zero-Shot).

| Model | Precision | Recall/Sensitivity | Specificity | F1 (present) | Macro-Average | Accuracy | BAC | MCC |
|---|---|---|---|---|---|---|---|---|
| Llama-3.1-8b | 0.763 | 0.827 | 0.411 | 0.794 | 0.624 | 0.701 | 0.619 | 0.254 |
| Llama-3.3-70b | 0.764 | 0.952 | 0.324 | 0.847 | 0.649 | 0.761 | 0.638 | 0.374 |
| Llama-OpenBio | 0.746 | 0.951 | 0.263 | 0.836 | 0.609 | 0.741 | 0.607 | 0.309 |
| Mistral-nemo | 0.731 | 0.922 | 0.222 | 0.816 | 0.566 | 0.710 | 0.572 | 0.202 |
| Mistral-small | 0.790 | 0.873 | 0.469 | 0.830 | 0.681 | 0.750 | 0.671 | 0.374 |
| Mistral-large | 0.765 | 0.941 | 0.335 | 0.844 | 0.650 | 0.757 | 0.638 | 0.362 |
| Gemma-3-27B | 0.744 | 0.953 | 0.249 | 0.836 | 0.601 | 0.739 | 0.601 | 0.299 |
| Gemma-3-27B (Med) | 0.741 | 0.967 | 0.226 | 0.839 | 0.593 | 0.741 | 0.597 | 0.308 |
| Qwen 3 Next Instruct | 0.759 | 0.939 | 0.316 | 0.840 | 0.637 | 0.750 | 0.628 | 0.340 |
| Qwen 3 Next Thinking | 0.774 | 0.918 | 0.383 | 0.839 | 0.663 | 0.755 | 0.650 | 0.365 |
| Qwen 3 A235B Instruct | 0.781 | 0.941 | 0.394 | 0.853 | 0.684 | 0.775 | 0.668 | 0.419 |
| Qwen 3 A235B Thinking | 0.768 | 0.922 | 0.362 | 0.838 | 0.654 | 0.752 | 0.642 | 0.353 |
| Deepseek-R1-Qwen | 0.766 | 0.928 | 0.347 | 0.839 | 0.649 | 0.752 | 0.638 | 0.350 |
| Deepseek-R1-Llama | 0.780 | 0.918 | 0.410 | 0.843 | 0.678 | 0.763 | 0.664 | 0.391 |

Supplementary Table 14. Full Large Language Model results (Zero-Shot) for detecting presence or absence of Hallucinations.

| Model | Precision | Recall/Sensitivity | Specificity | F1 (present) | Macro-Average | Accuracy | BAC | MCC |
|---|---|---|---|---|---|---|---|---|
| Llama-3.1-8b | 0.916 | 0.916 | 0.277 | 0.916 | 0.597 | 0.850 | 0.596 | 0.194 |
| Llama-3.3-70b | 0.922 | 0.959 | 0.301 | 0.940 | 0.652 | 0.891 | 0.630 | 0.315 |
| Llama-OpenBio | 0.902 | 0.988 | 0.099 | 0.943 | 0.554 | 0.894 | 0.543 | 0.187 |
| Mistral-nemo | 0.912 | 0.971 | 0.192 | 0.941 | 0.604 | 0.890 | 0.582 | 0.239 |
| Mistral-small | 0.958 | 0.851 | 0.676 | 0.902 | 0.677 | 0.833 | 0.764 | 0.397 |
| Mistral-large | 0.929 | 0.935 | 0.384 | 0.932 | 0.663 | 0.878 | 0.659 | 0.327 |
| Gemma-3-27B | 0.918 | 0.954 | 0.260 | 0.936 | 0.625 | 0.882 | 0.607 | 0.259 |
| Gemma-3-27B (Med) | 0.923 | 0.952 | 0.315 | 0.937 | 0.651 | 0.886 | 0.634 | 0.309 |
| Qwen 3 Next Instruct | 0.928 | 0.947 | 0.361 | 0.937 | 0.666 | 0.886 | 0.654 | 0.336 |
| Qwen 3 Next Thinking | 0.952 | 0.893 | 0.612 | 0.922 | 0.702 | 0.864 | 0.753 | 0.421 |
| Qwen 3 A235B Instruct | 0.943 | 0.938 | 0.507 | 0.940 | 0.719 | 0.893 | 0.723 | 0.437 |
| Qwen 3 A235B Thinking | 0.955 | 0.908 | 0.630 | 0.931 | 0.725 | 0.879 | 0.769 | 0.462 |
| Deepseek-R1-Qwen | 0.938 | 0.916 | 0.479 | 0.927 | 0.681 | 0.871 | 0.698 | 0.365 |
| Deepseek-R1-Llama | 0.939 | 0.907 | 0.498 | 0.923 | 0.678 | 0.864 | 0.702 | 0.362 |

Supplementary Table 15. Full Large Language Model results (Zero-Shot) for detecting presence or absence of Delusions.

| Model | Precision | Recall/Sensitivity | Specificity | F1 (present) | Macro-Average | Accuracy | BAC | MCC |
|---|---|---|---|---|---|---|---|---|
| Llama-3.1-8b | 0.480 | 0.705 | 0.563 | 0.571 | 0.611 | 0.615 | 0.634 | 0.259 |
| Llama-3.3-70b | 0.692 | 0.835 | 0.783 | 0.756 | 0.795 | 0.802 | 0.809 | 0.600 |
| Llama-OpenBio | 0.411 | 0.967 | 0.202 | 0.577 | 0.453 | 0.481 | 0.584 | 0.234 |
| Mistral-nemo | 0.543 | 0.793 | 0.611 | 0.644 | 0.675 | 0.678 | 0.702 | 0.391 |
| Mistral-small | 0.792 | 0.447 | 0.932 | 0.571 | 0.700 | 0.754 | 0.690 | 0.451 |
| Mistral-large | 0.741 | 0.775 | 0.842 | 0.758 | 0.805 | 0.817 | 0.808 | 0.611 |
| Gemma-3-27B | 0.576 | 0.884 | 0.621 | 0.697 | 0.716 | 0.718 | 0.752 | 0.491 |
| Gemma-3-27B (Med) | 0.546 | 0.863 | 0.582 | 0.669 | 0.685 | 0.685 | 0.722 | 0.435 |
| Qwen 3 Next Instruct | 0.610 | 0.880 | 0.672 | 0.720 | 0.746 | 0.748 | 0.776 | 0.533 |
| Qwen 3 Next Thinking | 0.670 | 0.827 | 0.762 | 0.740 | 0.779 | 0.786 | 0.795 | 0.571 |
| Qwen 3 A235B Instruct | 0.789 | 0.752 | 0.883 | 0.770 | 0.820 | 0.835 | 0.817 | 0.641 |
| Qwen 3 A235B Thinking | 0.698 | 0.814 | 0.795 | 0.751 | 0.793 | 0.802 | 0.804 | 0.593 |
| Deepseek-R1-Qwen | 0.749 | 0.764 | 0.851 | 0.756 | 0.806 | 0.819 | 0.807 | 0.612 |
| Deepseek-R1-Llama | 0.805 | 0.670 | 0.906 | 0.731 | 0.798 | 0.820 | 0.788 | 0.603 |

Supplementary Table 16. Full Large Language Model results (Zero-Shot) for detecting presence or absence of PFTD.

| Model | Precision | Recall/Sensitivity | Specificity | F1 (present) | Macro-Average | Accuracy | BAC | MCC |
|---|---|---|---|---|---|---|---|---|
| Llama-3.1-8b | 0.918 | 0.709 | 0.509 | 0.800 | 0.535 | 0.686 | 0.609 | 0.149 |
| Llama-3.3-70b | 0.930 | 0.850 | 0.500 | 0.888 | 0.631 | 0.810 | 0.675 | 0.283 |
| Llama-OpenBio | 0.897 | 0.979 | 0.138 | 0.936 | 0.574 | 0.882 | 0.558 | 0.203 |
| Mistral-nemo | 0.924 | 0.820 | 0.475 | 0.869 | 0.600 | 0.780 | 0.647 | 0.228 |
| Mistral-small | 0.989 | 0.283 | 0.975 | 0.440 | 0.349 | 0.362 | 0.629 | 0.189 |
| Mistral-large | 0.964 | 0.693 | 0.800 | 0.806 | 0.594 | 0.705 | 0.746 | 0.326 |
| Gemma-3-27B | 0.936 | 0.776 | 0.588 | 0.848 | 0.600 | 0.754 | 0.682 | 0.261 |
| Gemma-3-27B (Med) | 0.942 | 0.725 | 0.650 | 0.819 | 0.581 | 0.717 | 0.688 | 0.256 |
| Qwen 3 Next Instruct | 0.931 | 0.895 | 0.483 | 0.913 | 0.666 | 0.848 | 0.689 | 0.338 |
| Qwen 3 Next Thinking | 0.938 | 0.805 | 0.583 | 0.866 | 0.621 | 0.780 | 0.694 | 0.289 |
| Qwen 3 A235B Instruct | 0.938 | 0.827 | 0.575 | 0.879 | 0.636 | 0.798 | 0.701 | 0.309 |
| Qwen 3 A235B Thinking | 0.934 | 0.837 | 0.537 | 0.883 | 0.632 | 0.803 | 0.687 | 0.294 |
| Deepseek-R1-Qwen | 0.955 | 0.681 | 0.750 | 0.795 | 0.575 | 0.689 | 0.716 | 0.284 |
| Deepseek-R1-Llama | 0.959 | 0.618 | 0.796 | 0.752 | 0.544 | 0.639 | 0.707 | 0.266 |

Supplementary Table 17. Full Large Language Model results (Zero-Shot) for detecting presence or absence of Bizarre Behaviour.

| Model | Precision | Recall/Sensitivity | Specificity | F1 (present) | Macro-Average | Accuracy | BAC | MCC |
|---|---|---|---|---|---|---|---|---|
| Llama-3.1-8b | 0.594 | 0.532 | 0.598 | 0.561 | 0.563 | 0.563 | 0.565 | 0.130 |
| Llama-3.3-70b | 0.684 | 0.498 | 0.749 | 0.576 | 0.614 | 0.618 | 0.624 | 0.254 |
| Llama-OpenBio | 0.518 | 0.975 | 0.030 | 0.677 | 0.367 | 0.519 | 0.502 | 0.015 |
| Mistral-nemo | 0.608 | 0.427 | 0.701 | 0.502 | 0.553 | 0.559 | 0.564 | 0.133 |
| Mistral-small | 0.851 | 0.202 | 0.961 | 0.327 | 0.503 | 0.566 | 0.582 | 0.248 |
| Mistral-large | 0.677 | 0.478 | 0.753 | 0.560 | 0.605 | 0.610 | 0.615 | 0.239 |
| Gemma-3-27B | 0.699 | 0.475 | 0.777 | 0.566 | 0.614 | 0.620 | 0.626 | 0.263 |
| Gemma-3-27B (Med) | 0.632 | 0.577 | 0.633 | 0.603 | 0.603 | 0.603 | 0.605 | 0.209 |
| Qwen 3 Next Instruct | 0.673 | 0.522 | 0.724 | 0.588 | 0.617 | 0.619 | 0.623 | 0.250 |
| Qwen 3 Next Thinking | 0.693 | 0.481 | 0.768 | 0.568 | 0.613 | 0.618 | 0.624 | 0.258 |
| Qwen 3 A235B Instruct | 0.710 | 0.475 | 0.789 | 0.570 | 0.619 | 0.625 | 0.632 | 0.277 |
| Qwen 3 A235B Thinking | 0.700 | 0.511 | 0.762 | 0.591 | 0.627 | 0.631 | 0.636 | 0.281 |
| Deepseek-R1-Qwen | 0.749 | 0.440 | 0.839 | 0.554 | 0.620 | 0.631 | 0.640 | 0.303 |
| Deepseek-R1-Llama | 0.690 | 0.445 | 0.783 | 0.541 | 0.599 | 0.608 | 0.614 | 0.242 |

Supplementary Table 18. Full Large Language Model results (Zero-Shot) for detecting presence or absence of Affective Flattening.

| Model | Precision | Recall/Sensitivity | Specificity | F1 (present) | Macro-Average | Accuracy | BAC | MCC |
|---|---|---|---|---|---|---|---|---|
| Llama-3.1-8b | 0.433 | 0.616 | 0.404 | 0.509 | 0.494 | 0.494 | 0.510 | 0.020 |
| Llama-3.3-70b | 0.588 | 0.445 | 0.770 | 0.507 | 0.607 | 0.633 | 0.608 | 0.228 |
| Llama-OpenBio | 0.449 | 0.858 | 0.236 | 0.590 | 0.471 | 0.498 | 0.547 | 0.118 |
| Mistral-nemo | 0.458 | 0.478 | 0.590 | 0.467 | 0.534 | 0.543 | 0.534 | 0.068 |
| Mistral-small | 0.778 | 0.118 | 0.975 | 0.205 | 0.475 | 0.613 | 0.547 | 0.189 |
| Mistral-large | 0.618 | 0.326 | 0.851 | 0.426 | 0.575 | 0.628 | 0.588 | 0.209 |
| Gemma-3-27B | 0.581 | 0.587 | 0.689 | 0.584 | 0.638 | 0.646 | 0.638 | 0.276 |
| Gemma-3-27B (Med) | 0.538 | 0.617 | 0.609 | 0.575 | 0.609 | 0.613 | 0.613 | 0.224 |
| Qwen 3 Next Instruct | 0.590 | 0.509 | 0.740 | 0.547 | 0.625 | 0.642 | 0.624 | 0.255 |
| Qwen 3 Next Thinking | 0.554 | 0.529 | 0.687 | 0.541 | 0.608 | 0.620 | 0.608 | 0.217 |
| Qwen 3 A235B Instruct | 0.651 | 0.369 | 0.854 | 0.471 | 0.604 | 0.649 | 0.612 | 0.258 |
| Qwen 3 A235B Thinking | 0.599 | 0.487 | 0.759 | 0.537 | 0.624 | 0.643 | 0.623 | 0.256 |
| Deepseek-R1-Qwen | 0.660 | 0.352 | 0.867 | 0.460 | 0.600 | 0.649 | 0.610 | 0.259 |
| Deepseek-R1-Llama | 0.629 | 0.359 | 0.845 | 0.456 | 0.593 | 0.640 | 0.602 | 0.235 |

Supplementary Table 19. Full Large Language Model results (Zero-Shot) for detecting presence or absence of Alogia.

| Model | Precision | Recall/Sensitivity | Specificity | F1 (present) | Macro-Average | Accuracy | BAC | MCC |
|---|---|---|---|---|---|---|---|---|
| Llama-3.1-8b | 0.769 | 0.506 | 0.545 | 0.610 | 0.486 | 0.516 | 0.526 | 0.044 |
| Llama-3.3-70b | 0.794 | 0.565 | 0.574 | 0.660 | 0.532 | 0.567 | 0.569 | 0.121 |
| Llama-OpenBio | 0.750 | 0.943 | 0.085 | 0.836 | 0.486 | 0.724 | 0.514 | 0.050 |
| Mistral-nemo | 0.777 | 0.550 | 0.542 | 0.644 | 0.513 | 0.548 | 0.546 | 0.081 |
| Mistral-small | 0.795 | 0.119 | 0.910 | 0.208 | 0.307 | 0.321 | 0.515 | 0.041 |
| Mistral-large | 0.773 | 0.515 | 0.559 | 0.618 | 0.497 | 0.526 | 0.537 | 0.065 |
| Gemma-3-27B | 0.804 | 0.552 | 0.609 | 0.655 | 0.536 | 0.566 | 0.580 | 0.140 |
| Gemma-3-27B (Med) | 0.799 | 0.564 | 0.588 | 0.661 | 0.536 | 0.570 | 0.576 | 0.132 |
| Qwen 3 Next Instruct | 0.810 | 0.646 | 0.559 | 0.719 | 0.575 | 0.623 | 0.602 | 0.181 |
| Qwen 3 Next Thinking | 0.786 | 0.694 | 0.449 | 0.737 | 0.560 | 0.631 | 0.571 | 0.131 |
| Qwen 3 A235B Instruct | 0.792 | 0.592 | 0.547 | 0.678 | 0.539 | 0.581 | 0.570 | 0.122 |
| Qwen 3 A235B Thinking | 0.803 | 0.607 | 0.564 | 0.691 | 0.554 | 0.596 | 0.586 | 0.151 |
| Deepseek-R1-Qwen | 0.782 | 0.364 | 0.704 | 0.497 | 0.446 | 0.451 | 0.534 | 0.062 |
| Deepseek-R1-Llama | 0.809 | 0.456 | 0.684 | 0.583 | 0.500 | 0.514 | 0.570 | 0.124 |

Supplementary Table 20. Full Large Language Model results (Zero-Shot) for detecting presence or absence of Anhedonia & Asociality.

| Model | Precision | Recall/Sensitivity | Specificity | F1 (present) | Macro-Average | Accuracy | BAC | MCC |
|---|---|---|---|---|---|---|---|---|
| Llama-3.1-8b | 0.822 | 0.516 | 0.566 | 0.634 | 0.482 | 0.527 | 0.541 | 0.067 |
| Llama-3.3-70b | 0.849 | 0.595 | 0.593 | 0.699 | 0.538 | 0.594 | 0.594 | 0.153 |
| Llama-OpenBio | 0.803 | 0.958 | 0.103 | 0.874 | 0.519 | 0.780 | 0.531 | 0.110 |
| Mistral-nemo | 0.854 | 0.410 | 0.731 | 0.554 | 0.460 | 0.476 | 0.571 | 0.118 |
| Mistral-small | 0.896 | 0.078 | 0.966 | 0.143 | 0.247 | 0.262 | 0.522 | 0.069 |
| Mistral-large | 0.866 | 0.508 | 0.697 | 0.640 | 0.514 | 0.547 | 0.602 | 0.166 |
| Gemma-3-27B | 0.859 | 0.621 | 0.607 | 0.721 | 0.558 | 0.618 | 0.614 | 0.186 |
| Gemma-3-27B (Med) | 0.836 | 0.622 | 0.531 | 0.713 | 0.535 | 0.603 | 0.577 | 0.126 |
| Qwen 3 Next Instruct | 0.855 | 0.657 | 0.572 | 0.743 | 0.569 | 0.639 | 0.614 | 0.190 |
| Qwen 3 Next Thinking | 0.842 | 0.689 | 0.501 | 0.758 | 0.564 | 0.650 | 0.595 | 0.161 |
| Qwen 3 A235B Instruct | 0.861 | 0.589 | 0.634 | 0.699 | 0.547 | 0.598 | 0.612 | 0.181 |
| Qwen 3 A235B Thinking | 0.866 | 0.646 | 0.614 | 0.740 | 0.576 | 0.639 | 0.630 | 0.214 |
| Deepseek-R1-Qwen | 0.861 | 0.478 | 0.703 | 0.614 | 0.496 | 0.524 | 0.591 | 0.148 |
| Deepseek-R1-Llama | 0.861 | 0.552 | 0.660 | 0.673 | 0.532 | 0.575 | 0.606 | 0.172 |

Supplementary Table 21. Full Large Language Model results (Zero-Shot) for detecting presence or absence of Avolition & Apathy.

#### Full tables of results for the validation analyses.

| | **Hallucinations** | | | | **Delusions** | | | | **PFTD** | | | | **Bizarre Behaviour** | | | |
|---|---|---|---|---|---|---|---|---|---|---|---|---|---|---|---|---|
| **Variable** | **OR** | **95% CI** | **p value** | **FDR** | **OR** | **95% CI** | **p value** | **FDR** | **OR** | **95% CI** | **p value** | **FDR** | **OR** | **95% CI** | **p value** | **FDR** |
| Age of Onset | 0.972 | 0.945 - 0.999 | **0.041** | 0.066 | 1.029 | 0.995 - 1.066 | 0.099 | 0.132 | 0.982 | 0.962 - 1.002 | 0.074 | 0.148 | 0.998 | 0.974 - 1.022 | 0.848 | 0.848 |
| Disorder Course | 1.947 | 1.535 - 2.484 | **5x10$^{-8}$** | **4x10$^{-7}$** | 0.970 | 0.746 - 1.249 | 0.813 | 0.813 | 0.964 | 0.814 - 1.141 | 0.665 | 0.761 | 1.252 | 1.016 - 1.54 | **0.034** | 0.09 |
| Clozapine Use | 2.384 | 1.409 - 4.206 | **0.002** | **0.005** | 1.963 | 1.144 - 3.501 | **0.017** | **0.035** | 0.893 | 0.643 - 1.238 | 0.499 | 0.665 | 1.419 | 0.928 - 2.2 | 0.111 | 0.177 |
| GAS (Worst in Psychosis) | 0.938 | 0.912 - 0.965 | **7x10$^{-6}$** | **3x10$^{-5}$** | 0.922 | 0.894 - 0.95 | **1x10$^{-7}$** | **1x10$^{-6}$** | 0.930 | 0.908 - 0.951 | **8x10$^{-10}$** | **7x10$^{-9}$** | 0.942 | 0.918 - 0.965 | **2x10$^{-6}$** | **2x10$^{-5}$** |
| Depot Use | 1.467 | 0.914 - 2.364 | 0.113 | 0.151 | 1.633 | 0.972 - 2.776 | 0.066 | 0.106 | 1.395 | 1.002 - 1.947 | **0.049** | 0.131 | 2.257 | 1.474 - 3.487 | **2x10$^{-4}$** | **8x10$^{-4}$** |
| Sex (F) | 1.347 | 0.837 - 2.204 | 0.226 | 0.259 | 0.800 | 0.487 - 1.317 | 0.376 | 0.43 | 0.955 | 0.686 - 1.329 | 0.786 | 0.786 | 0.767 | 0.509 - 1.159 | 0.205 | 0.274 |
| Age at Interview | 0.989 | 0.971 - 1.008 | 0.273 | 0.273 | 1.027 | 1.005 - 1.049 | **0.015** | **0.035** | 0.991 | 0.977 - 1.004 | 0.167 | 0.267 | 0.984 | 0.968 - 1.001 | 0.062 | 0.124 |
| Schizophrenia diagnosis | 1.843 | 1.153 - 2.94 | **0.01** | **0.02** | 2.560 | 1.561 - 4.227 | **2x10$^{-4}$** | **8x10$^{-4}$** | 1.405 | 1.003 - 1.975 | **0.049** | 0.131 | 1.268 | 0.834 - 1.918 | 0.263 | 0.301 |

Supplementary Table 22. Associations with LLM-predicted positive symptoms. All results are from univariate GLMs controlling for diagnosis, age and sex, except for diagnosis, age, and sex which each covary for each other. OR = Odds Ratio; CI = 95% Confidence Intervals.

| | Affective Flattening | | | | Alogia | | | | Anhedonia & Asociality | | | | Avolition & Apathy | | | |
|---|---|---|---|---|---|---|---|---|---|---|---|---|---|---|---|---|
| **Variable** | **OR** | **95% CI** | **p value** | **FDR** | **OR** | **95% CI** | **p value** | **FDR** | **OR** | **95% CI** | **p value** | **FDR** | **OR** | **95% CI** | **p value** | **FDR** |
| Age of Onset | 0.982 | 0.962 - 1.002 | 0.081 | 0.16 | 0.975 | 0.955 - 0.995 | **0.018** | 0.073 | 0.993 | 0.974 - 1.013 | 0.482 | 0.642 | 0.991 | 0.972 - 1.011 | 0.383 | 0.588 |
| Disorder Course | 1.276 | 1.071 - 1.525 | **0.007** | **0.027** | 1.016 | 0.854 - 1.211 | 0.856 | 0.856 | 1.171 | 0.988 - 1.388 | 0.069 | 0.274 | 1.187 | 1.001 - 1.409 | **0.049** | 0.148 |
| Clozapine Use | 1.319 | 0.948 - 1.835 | 0.1 | 0.16 | 1.227 | 0.875 - 1.719 | 0.234 | 0.375 | 1.142 | 0.824 - 1.586 | 0.425 | 0.642 | 0.829 | 0.595 - 1.155 | 0.268 | 0.535 |
| GAS (Worst in Psychosis) | 0.960 | 0.939 - 0.981 | **$3x10^{-4}$** | **0.002** | 0.966 | 0.945 - 0.987 | **0.002** | **0.016** | 0.997 | 0.977 - 1.017 | 0.755 | 0.772 | 0.980 | 0.96 - 1 | 0.056 | 0.148 |
| Depot Use | 1.366 | 0.977 - 1.916 | 0.069 | 0.16 | 1.259 | 0.894 - 1.778 | 0.189 | 0.375 | 0.858 | 0.616 - 1.194 | 0.365 | 0.642 | 1.044 | 0.746 - 1.46 | 0.801 | 0.801 |
| Sex (F) | 0.913 | 0.652 - 1.276 | 0.596 | 0.681 | 0.889 | 0.63 - 1.25 | 0.499 | 0.612 | 0.832 | 0.598 - 1.157 | 0.274 | 0.642 | 0.702 | 0.503 - 0.979 | **0.037** | 0.148 |
| Age at Interview | 0.994 | 0.98 - 1.007 | 0.347 | 0.463 | 0.996 | 0.982 - 1.009 | 0.535 | 0.612 | 1.002 | 0.989 - 1.015 | 0.772 | 0.772 | 0.995 | 0.981 - 1.008 | 0.441 | 0.588 |
| Schizophrenia diagnosis | 1.021 | 0.726 - 1.439 | 0.905 | 0.905 | 0.808 | 0.572 - 1.143 | 0.227 | 0.375 | 0.692 | 0.492 - 0.969 | **0.033** | 0.265 | 0.947 | 0.672 - 1.329 | 0.754 | 0.801 |

Supplementary Table 23. Associations with LLM-predicted negative symptoms. All results are from univariate GLMs controlling for diagnosis, age and sex, except for diagnosis, age, and sex which each covary for each other. OR = Odds Ratio; CI = 95% Confidence Intervals.

| | Hallucinations | | | | Delusions | | | | PFTD | | | | Bizarre Behaviour | | | |
|---|---|---|---|---|---|---|---|---|---|---|---|---|---|---|---|---|
| **Variable** | **OR** | **95% CI** | **p value** | **FDR** | **OR** | **95% CI** | **p value** | **FDR** | **OR** | **95% CI** | **p value** | **FDR** | **OR** | **95% CI** | **p value** | **FDR** |
| Age of Onset | 0.956 | 0.934 - 0.977 | **$6x10^{-5}$** | **$5x10^{-4}$** | 0.987 | 0.95 - 1.025 | 0.495 | 0.66 | 0.968 | 0.947 - 0.988 | **0.002** | **0.006** | 0.998 | 0.967 - 1.03 | 0.897 | 0.897 |
| Disorder Course | 1.406 | 1.169 - 1.694 | **$3x10^{-4}$** | **0.001** | 1.249 | 0.908 - 1.706 | 0.165 | 0.331 | 1.149 | 0.965 - 1.372 | 0.122 | 0.196 | 1.144 | 0.851 - 1.522 | 0.363 | 0.58 |
| Clozapine Use | 1.567 | 1.08 - 2.294 | **0.019** | **0.031** | 1.531 | 0.8 - 3.095 | 0.213 | 0.341 | 1.050 | 0.751 - 1.466 | 0.773 | 0.773 | 1.154 | 0.653 - 2.091 | 0.628 | 0.736 |
| GAS (Worst in Psychosis) | 0.968 | 0.947 - 0.99 | **0.004** | **0.011** | 0.920 | 0.888 - 0.953 | **$4x10^{-6}$** | **$3x10^{-5}$** | 0.936 | 0.914 - 0.958 | **$5x10^{-8}$** | **$4x10^{-7}$** | 0.927 | 0.897 - 0.957 | **$5x10^{-6}$** | **$4x10^{-5}$** |
| Depot Use | 1.618 | 1.121 - 2.343 | **0.01** | **0.021** | 1.622 | 0.87 - 3.081 | 0.132 | 0.331 | 1.788 | 1.27 - 2.527 | **$9x10^{-4}$** | **0.004** | 1.768 | 1.002 - 3.153 | 0.05 | 0.134 |
| Sex (F) | 1.274 | 0.883 - 1.852 | 0.2 | 0.2 | 0.884 | 0.482 - 1.637 | 0.693 | 0.792 | 0.870 | 0.618 - 1.222 | 0.423 | 0.564 | 1.145 | 0.651 - 2.057 | 0.644 | 0.736 |
| Age at Interview | 0.989 | 0.975 - 1.004 | 0.144 | 0.165 | 0.998 | 0.974 - 1.024 | 0.899 | 0.899 | 0.996 | 0.982 - 1.01 | 0.564 | 0.645 | 0.976 | 0.954 - 0.998 | **0.037** | 0.134 |
| Schizophrenia diagnosis | 1.525 | 1.056 - 2.199 | **0.024** | **0.032** | 2.794 | 1.52 - 5.225 | **0.001** | **0.004** | 1.549 | 1.092 - 2.209 | **0.015** | **0.03** | 1.394 | 0.788 - 2.44 | 0.248 | 0.495 |

Supplementary Table 24. Associations with Gold Standard positive symptoms. All results are from univariate GLMs controlling for diagnosis, age and sex, except for diagnosis, age, and sex which each covary for each other. OR = Odds Ratio; CI = 95% Confidence Intervals.

| | Affective Flattening | | | | Alogia | | | | Anhedonia & Asociality | | | | Avolition & Apathy | | | |
|---|---|---|---|---|---|---|---|---|---|---|---|---|---|---|---|---|
| **Variable** | **OR** | **95% CI** | **p value** | **FDR** | **OR** | **95% CI** | **p value** | **FDR** | **OR** | **95% CI** | **p value** | **FDR** | **OR** | **95% CI** | **p value** | **FDR** |
| Age of Onset | 0.993 | 0.973 - 1.012 | 0.452 | 0.603 | 0.996 | 0.977 - 1.016 | 0.725 | 0.725 | 0.983 | 0.961 - 1.006 | 0.136 | 0.211 | 0.957 | 0.934 - 0.981 | **$5x10^{-4}$** | **0.002** |
| Disorder Course | 1.488 | 1.253 - 1.774 | **$7x10^{-6}$** | **$6x10^{-5}$** | 1.102 | 0.931 - 1.308 | 0.262 | 0.349 | 1.678 | 1.388 - 2.034 | **$1x10^{-7}$** | **$8x10^{-7}$** | 2.105 | 1.716 - 2.598 | **$2x10^{-12}$** | **$1x10^{-11}$** |
| Clozapine Use | 1.356 | 0.98 - 1.881 | 0.066 | 0.177 | 1.282 | 0.925 - 1.777 | 0.135 | 0.271 | 1.296 | 0.887 - 1.909 | 0.184 | 0.211 | 1.825 | 1.202 - 2.817 | **0.006** | **0.011** |
| GAS (Worst in Psychosis) | 0.989 | 0.97 - 1.009 | 0.297 | 0.594 | 0.972 | 0.951 - 0.992 | 0.007 | 0.057 | 1.017 | 0.993 - 1.042 | 0.168 | 0.211 | 0.959 | 0.937 - 0.983 | **$7x10^{-4}$** | **0.002** |
| Depot Use | 1.150 | 0.829 - 1.597 | 0.401 | 0.603 | 1.410 | 1.011 - 1.97 | **0.043** | 0.173 | 1.045 | 0.713 - 1.529 | 0.82 | 0.82 | 1.614 | 1.079 - 2.421 | **0.02** | **0.032** |
| Sex (F) | 0.731 | 0.526 - 1.013 | 0.06 | 0.177 | 0.734 | 0.525 - 1.022 | 0.068 | 0.181 | 0.747 | 0.515 - 1.086 | 0.125 | 0.211 | 0.900 | 0.605 - 1.346 | 0.606 | 0.69 |
| Age at Interview | 0.998 | 0.985 - 1.012 | 0.808 | 0.912 | 0.995 | 0.982 - 1.008 | 0.455 | 0.52 | 1.012 | 0.997 - 1.028 | 0.127 | 0.211 | 1.003 | 0.987 - 1.02 | 0.69 | 0.69 |
| Schizophrenia diagnosis | 1.019 | 0.73 - 1.422 | 0.912 | 0.912 | 1.221 | 0.871 - 1.715 | 0.247 | 0.349 | 1.362 | 0.932 - 1.986 | 0.109 | 0.211 | 1.256 | 0.837 - 1.875 | 0.267 | 0.356 |

Supplementary Table 25. Associations with Gold Standard negative symptoms. All results are from univariate GLMs controlling for diagnosis, age and sex, except for diagnosis, age, and sex which each covary for each other. OR = Odds Ratio; CI = 95% Confidence Intervals.

Full tables of results for the error analysis.

| **Variable** | **B** | **SE** | **p value** |
|---|---|---|---|
| log(N) | 0.113 | 0.023 | **$1x10^{-6}$** |
| Sex2 | -0.020 | 0.033 | 0.545 |
| Age_at_Interview | -0.001 | 0.001 | 0.435 |
| Ethnic_Origin | 0.022 | 0.083 | 0.787 |
| ICD_diagnosis_new | -0.040 | 0.064 | 0.531 |
| sumstratS2a | 0.055 | 0.206 | 0.787 |
| sumstratS2b | 0.005 | 0.231 | 0.983 |
| sumstratS2c | 0.197 | 0.148 | 0.182 |
| sumstratS2d | 0.096 | 0.047 | **0.042** |
| sumstratS2e | -0.032 | 0.184 | 0.863 |
| sumstratS2f | -0.061 | 0.290 | 0.834 |
| sumstratS2g | -0.006 | 0.073 | 0.940 |
| sumstratUnknown | 0.013 | 0.072 | 0.856 |

Supplementary Table 26. Full regression results assessing effect of descriptive variables on the number of correctly predicted symptoms made by Qwen3 235B Thinking. Sumstrat groupings represent groups of case note summarisers.

| Error | Definition | Hallucinations | Delusions | PFTD | Bizarre Behaviour | Affective Flattening | Alogia | Anhedonia & Asociality | Avolition & Apathy |
|---|---|---|---|---|---|---|---|---|---|
| Hyper-sensitivity | Little, vague, or limited evidence for the symptom in the case note that would not be at an appropriate level for a positive rating. | 12 (60%) | 4 (20%) | 13 (65%) | 4 (20%) | 6 (30%) | 4 (20%) | 2 (10%) | 4 (20%) |
| Hypo-sensitivity | Evidence for the symptom exists in the case note that the model has not picked up on or discounted. | 1 (5%) | 3 (15%) | 0 (0%) | 0 (0%) | 0 (0%) | 0 (0%) | 1 (5%) | 0 (0%) |
| Information Conflicting | Evidence exists both for and against the symptom's presence/absence in the case note, with the model prioritising information leading to the incorrect rating. | 3 (15%) | 1 (5%) | 0 (0%) | 0 (0%) | 0 (0%) | 0 (0%) | 1 (5%) | 1 (5%) |
| Information Limited | There is no evidence in the case note that would support a positive rating being made. This has been correctly identified by the model. | 6 (30%) | 14 (70%) | 7 (35%) | 13 (65%) | 13 (65%) | 13 (65%) | 17 (85%) | 16 (80%) |
| Symptom Misattribution | There is evidence for another symptom that is mistakenly taken as evidence for the symptom of interest. Alternatively, a symptom present but its due to the effects of drugs or otherwise rather than psychiatric disorder | 2 (10%) | 2 (10%) | 0 (0%) | 3 (15%) | 1 (5%) | 3 (15%) | 0 (0%) | 0 (0%) |
| Definition Error | Model's reasoning is made on incorrect assumptions about the symptom in question. | 0 (0%) | 0 (0%) | 0 (0%) | 0 (0%) | 1 (5%) | 3 (15%) | 0 (0%) | 0 (0%) |

Supplementary Table 27. Full breakdown of errors identified in error analysis and their definitions.

| Symptom | Total Assessed | Total Recoded (%) |
|---|---|---|
| Hallucinations | 130 | 81 (62.3%) |
| Delusions | 26 | 17 (65.4%) |
| PFTD | 74 | 17 (23%) |
| Bizarre Behaviour | 16 | 10 (62.5%) |
| Affective Flattening | 40 | 10 (25%) |
| Alogia | 34 | 7 (20.6%) |
| Anhedonia & Asociality | 41 | 12 (40 %) |
| Avolition & Apathy | 30 | 12 (29.3%) |

Supplementary Table 28. Changes in gold-standard symptom classifications after reclassification using solely clinical summary information.

#### Tables of results for few-shot prompts and Large Language Models (LLMs)

| Model | Precision | Recall/Sensitivity | Specificity | F1 (present) | Macro-Average | Accuracy | BAC | MCC |
|---|---|---|---|---|---|---|---|---|
| Deepseek-R1-Qwen | 0.805 | 0.614 | 0.700 | 0.682 | 0.611 | 0.671 | 0.657 | 0.298 |
| Qwen 3 A235B Thinking | 0.797 | 0.710 | 0.643 | 0.743 | 0.648 | 0.717 | 0.677 | 0.332 |
| Qwen 3 Next Thinking | 0.775 | 0.724 | 0.597 | 0.741 | 0.635 | 0.709 | 0.660 | 0.299 |

Supplementary Table 29. Comparing Mean Metrics across symptoms for Large Language Models (Few-Shot).

| Model | Precision | Recall/Sensitivity | Specificity | F1 (present) | Macro-Average | Accuracy | BAC | MCC |
|---|---|---|---|---|---|---|---|---|
| Qwen 3 A235B Thinking | 0.772 | 0.920 | 0.376 | 0.840 | 0.661 | 0.755 | 0.648 | 0.362 |
| Qwen 3 Next Thinking | 0.768 | 0.912 | 0.366 | 0.834 | 0.650 | 0.746 | 0.639 | 0.339 |
| Deepseek-R1-Qwen | 0.774 | 0.922 | 0.380 | 0.841 | 0.665 | 0.758 | 0.651 | 0.371 |

Supplementary Table 30. Full LLM results (Few-Shot) for detecting presence or absence of Hallucinations.

| Model | Precision | Recall/Sensitivity | Specificity | F1 (present) | Macro-Average | Accuracy | BAC | MCC |
|---|---|---|---|---|---|---|---|---|
| Qwen 3 A235B Thinking | 0.951 | 0.898 | 0.603 | 0.924 | 0.705 | 0.868 | 0.751 | 0.424 |
| Qwen 3 Next Thinking | 0.953 | 0.899 | 0.616 | 0.925 | 0.710 | 0.869 | 0.758 | 0.434 |
| Deepseek-R1-Qwen | 0.947 | 0.900 | 0.562 | 0.923 | 0.693 | 0.865 | 0.731 | 0.397 |

Supplementary Table 31. Full LLM results (Few-Shot) for detecting presence or absence of Delusions.

| Model | Precision | Recall/Sensitivity | Specificity | F1 (present) | Macro-Average | Accuracy | BAC | MCC |
|---|---|---|---|---|---|---|---|---|
| Qwen 3 A235B Thinking | 0.692 | 0.845 | 0.781 | 0.761 | 0.798 | 0.805 | 0.813 | 0.607 |
| Qwen 3 Next Thinking | 0.627 | 0.841 | 0.709 | 0.719 | 0.753 | 0.757 | 0.775 | 0.530 |
| Deepseek-R1-Qwen | 0.745 | 0.748 | 0.851 | 0.747 | 0.799 | 0.813 | 0.800 | 0.599 |

Supplementary Table 32. Full LLM results (Few-Shot) for detecting presence or absence of Positive Formal Thought Disorder.

| Model | Precision | Recall/Sensitivity | Specificity | F1 (present) | Macro-Average | Accuracy | BAC | MCC |
|---|---|---|---|---|---|---|---|---|
| Qwen 3 A235B Thinking | 0.942 | 0.814 | 0.613 | 0.873 | 0.637 | 0.791 | 0.713 | 0.319 |
| Qwen 3 Next Thinking | 0.938 | 0.793 | 0.588 | 0.859 | 0.613 | 0.770 | 0.690 | 0.279 |
| Deepseek-R1-Qwen | 0.956 | 0.633 | 0.775 | 0.762 | 0.548 | 0.649 | 0.704 | 0.263 |

Supplementary Table 33. Full LLM results (Few-Shot) for detecting presence or absence of Bizarre Behaviour.

| Model | Precision | Recall/Sensitivity | Specificity | F1 (present) | Macro-Average | Accuracy | BAC | MCC |
|---|---|---|---|---|---|---|---|---|
| Qwen 3 A235B Thinking | 0.709 | 0.486 | 0.783 | 0.577 | 0.623 | 0.628 | 0.635 | 0.280 |
| Qwen 3 Next Thinking | 0.697 | 0.478 | 0.774 | 0.567 | 0.614 | 0.620 | 0.626 | 0.263 |
| Deepseek-R1-Qwen | 0.727 | 0.429 | 0.824 | 0.540 | 0.607 | 0.618 | 0.627 | 0.274 |

Supplementary Table 34. Full LLM results (Few-Shot) for detecting presence or absence of Affective Flattening.

| Model | Precision | Recall/Sensitivity | Specificity | F1 (present) | Macro-Average | Accuracy | BAC | MCC |
|---|---|---|---|---|---|---|---|---|
| Qwen 3 A235B Thinking | 0.637 | 0.478 | 0.800 | 0.546 | 0.640 | 0.664 | 0.639 | 0.295 |
| Qwen 3 Next Thinking | 0.579 | 0.493 | 0.736 | 0.533 | 0.615 | 0.633 | 0.615 | 0.236 |
| Deepseek-R1-Qwen | 0.649 | 0.372 | 0.852 | 0.473 | 0.605 | 0.649 | 0.612 | 0.258 |

Supplementary Table 35. Full LLM results (Few-Shot) for detecting presence or absence of Alogia.

| Model | Precision | Recall/Sensitivity | Specificity | F1 (present) | Macro-Average | Accuracy | BAC | MCC |
|---|---|---|---|---|---|---|---|---|
| Qwen 3 A235B Thinking | 0.800 | 0.634 | 0.536 | 0.707 | 0.560 | 0.609 | 0.585 | 0.151 |
| Qwen 3 Next Thinking | 0.771 | 0.709 | 0.385 | 0.739 | 0.542 | 0.626 | 0.547 | 0.088 |
| Deepseek-R1-Qwen | 0.793 | 0.411 | 0.687 | 0.541 | 0.473 | 0.481 | 0.549 | 0.088 |

Supplementary Table 36. Full LLM results (Few-Shot) for detecting presence or absence of Anhedonia & Asociality.

| Model | Precision | Recall/Sensitivity | Specificity | F1 (present) | Macro-Average | Accuracy | BAC | MCC |
|---|---|---|---|---|---|---|---|---|
| Qwen 3 A235B Thinking | 0.871 | 0.606 | 0.655 | 0.715 | 0.564 | 0.616 | 0.631 | 0.213 |
| Qwen 3 Next Thinking | 0.865 | 0.665 | 0.600 | 0.752 | 0.584 | 0.652 | 0.633 | 0.220 |
| Deepseek-R1-Qwen | 0.852 | 0.496 | 0.669 | 0.627 | 0.498 | 0.531 | 0.582 | 0.133 |

Supplementary Table 37. Full LLM results (Few-Shot) for detecting presence or absence of Avolition & Apathy.

#### Comparing performance across zero- and few-shot prompts

| **Symptom** | **McNemar's x2** | **p value** | **FDR *p* value** | **Best Prompt** | **Model** |
|---|---|---|---|---|---|
| Hallucinations | 0.727 | 0.394 | 0.736 | Zero | Deepseek Qwen 32B |
| Delusions | 0.364 | 0.546 | 0.736 | Zero | Deepseek Qwen 32B |
| PFTD | 0.258 | 0.611 | 0.736 | Zero | Deepseek Qwen 32B |
| Bizarre Behaviour | 5.939 | **0.015** | 0.118 | Zero | Deepseek Qwen 32B |
| Affective Flattening | 1.209 | 0.272 | 0.724 | Few | Deepseek Qwen 32B |
| Alogia | 0.000 | 1.000 | 1.000 | Few | Deepseek Qwen 32B |
| Avolition & Apathy | 0.214 | 0.644 | 0.736 | Few | Deepseek Qwen 32B |
| Anhedonia & Asociality | 3.645 | 0.056 | 0.225 | Few | Deepseek Qwen 32B |
| Hallucinations | 2.286 | 0.131 | 0.477 | Few | Qwen3 Next 80B |
| Delusions | 0.000 | 1.000 | 1.000 | Few | Qwen3 Next 80B |
| PFTD | 2.722 | 0.099 | 0.477 | Few | Qwen3 Next 80B |
| Bizarre Behaviour | 1.389 | 0.239 | 0.477 | Few | Qwen3 Next 80B |
| Affective Flattening | 0.026 | 0.873 | 0.997 | Zero | Qwen3 Next 80B |
| Alogia | 1.667 | 0.197 | 0.477 | Zero | Qwen3 Next 80B |
| Avolition & Apathy | 0.450 | 0.502 | 0.670 | Zero | Qwen3 Next 80B |
| Anhedonia & Asociality | 0.556 | 0.456 | 0.670 | Few | Qwen3 Next 80B |
| Hallucinations | 0.286 | 0.593 | 0.773 | Zero | Qwen3 235B |
| Delusions | 3.200 | 0.074 | 0.196 | Few | Qwen3 235B |
| PFTD | 0.083 | 0.773 | 0.773 | Few | Qwen3 235B |
| Bizarre Behaviour | 0.941 | 0.332 | 0.531 | Zero | Qwen3 235B |
| Affective Flattening | 0.125 | 0.724 | 0.773 | Zero | Qwen3 235B |
| Alogia | 4.900 | **0.027** | 0.118 | Zero | Qwen3 235B |
| Avolition & Apathy | 4.741 | **0.029** | 0.118 | Zero | Qwen3 235B |
| Anhedonia & Asociality | 1.141 | 0.285 | 0.531 | Few | Qwen3 235B |

Supplementary Table 38. McNemar's test comparing Zero- and Few-Shot performance across Symptoms and Models.

#### Guidelines for Reporting Machine Learning Investigations in Neuropsychiatry (GREMLIN)

| Section | Item # | Checklist item | Explanation | Complete |
|---|---|---|---|---|
| **TITLE** | | | | |
| Title | 1 | Identify the use of machine learning for predictive or diagnostic purposes. | This will allow more efficient database searching and identification of relevant studies. | |
| **ABSTRACT** | | | | |
| Article summary | 2 | Structured summary including background, aims, source data, model performance and conclusion. | The structured summary provides sufficient information for readers to grasp the range and main aspects of the study. | Pg1 |
| **INTRODUCTION** | | | | |
| Background | 3 | Describe the current knowledgebase and how learning algorithms may lead to improved patient care or mechanistic insights. | Enable the reader to follow the process of developing the research question; from what is known and clinically utilised in psychiatry, to the application and anticipated benefits of machine learning algorithms. | Pg2-3 |
| Aims | 4 | Describe the purpose of ML in terms of hypothesis testing or prediction. | Model interpretability is critical for hypotheses testing. Model interpretability is not critical for prediction purposes. | Pg2-3 |

| | | | | |
|---|---|---|---|---|
| | 5 | Predictive modelling: prognostic or diagnostic? | Predictive modelling may be prognostic (time to an event) or diagnostic (correctly identifying that a condition already exists). | Pg2-3 |
| | 6 | Frame the research question with respect to PICOS | PICOS: Population, intervention, comparator, outcome, study design.<br>More information regarding PICOS can be found in most institutional library guides. | |
| **METHODS** | | | | |
| Study design | 7 | Provide a full description, or reference to a full description of the population upon which modelling will occur; facility, screening dates and numbers, inclusion/exclusion criteria, database/study name, study duration. | Is the patient population from a prospective or retrospective study? For retrospective studies (where recruitment and data collection have already been performed), the reader must be able to gauge the fitness of the study for the current application. | Pg3-4 |
| | 8 | Include ethics statement and approval number | | Pg3-4 |
| Prediction environment | 9 | Describe the outcome measurement. | Outcome measurement(s) may include prediction per patient or per event. | Pg4-5 |

| | | | | |
|---|---|---|---|---|
| | 10 | Classification, regression or survival prediction | Classification is the prediction of a categorical label (such as good outcome or poor outcome).<br><br>Regression is the prediction of a quantity or continuous variable (such as score for depression).<br><br>Survival prediction is time to event and requires identification of the event (such as remission) and time measurement (such as weeks). | Pg4 |
| | 11 | Training and validation environments | Describe the process for model training and validation. This must include how data was partitioned. For example, how many patient samples contributed data for training? Was cross-validation used? How many folds? | Pg5 |
| | 12 | | Describe the process for hyperparameter optimisation. What parameters were tuned (eg Gamma, C), how were they tuned (eg grid search, random search or Bayes optimisation) and what ranges were used? Which optimisation criterion was used (eg prognostic summary index). Was model complexity accounted for? | Pg5 and Supplementary Materials |
| | 13 | Model testing | Model testing must be performed on new/unseen data. Describe the metrics to be used for assessment of model performance. For example sensitivity, specificity, positive predictive value, negative predictive | Pg7 |

| | | | | |
|---|---|---|---|---|
| | | | value, diagnostic odds ratio, Area Under the (receiver operating characteristic) Curve (AUC). | |
| | 14 | Data leakage | Data leakage may occur in relation to outcomes or validation and may lead to inflated prediction performance. The use of cross-validation, holding back data (unseen) for later validation and the use of pipeline architecture can reduce bias in model performance. Furthermore, data transformations should be performed separately on data partitions and not the entire data set to avoid leaking feature distribution information between folds. | Pg4-5 |
| | 15 | Prediction success | How will the model be determined as adequate for its purpose and how does it compare to other models? Model adequacy may include measures of discrimination and calibration. What statistical procedures were used for model comparison?<br><br>Global measures of diagnostic accuracy, such as Area Under Receiver Operating Characteristic Curve (AUC) allow for model comparison. | Pg7 |
| | 16 | Overfitting | Overfitting occurs when learning on the training data set is excessive and irrelevant or noisy features are included in selection leading to inflated model performance on training data but poor performance on | NA |

| | | | | |
|---|---|---|---|---|
| | | | unseen data. This can be overcome with k-fold cross validation and hold back validation data. | |
| Data acquisition and pre-processing | 17 | Present characteristics of the dataset | Provide relevant summary statistics for the dataset and particular information for the distribution or ratios of response variables. | Pg8 |
| | 18 | Explain the process for handling missing values, class imbalance, covariates and outliers | Clearly state where data were discarded or imputed (for example variables with >=20% missing values excluded). Manage class imbalance at data or algorithm level, or a combination of these. | Pg3 |
| | 19 | Check data for perfect separators | The data should be checked for uncommon values for categorical variables that may lead to overfitting. This can be overcome by removing the variable from selected features and assessing the impact. | NA |
| | 20 | Data scaling | What method was used for data scaling?<br>For example Z-score normalisation or Min-Max scaling (0-1). | NA |
| Outcomes, variables | 21 | Predictor variables | Describe predictor variables. | Pg7 |
| | 22 | Generalisability | Identify how generalisability will be addressed. For example, cross-validation, leave site out validation. | NA |

| | | | | |
|---|---|---|---|---|
| | 23 | Code/algorithm | Where in-house code is developed, it should be made available (for example deposited on Github), with appropriate commentary, to allow inspection.<br><br>Where a code package was used (for example R package e1071, Python package Scikit-learn and NeuroMiner) this should be stated. | Pg5 |
| **RESULTS** | | | | |
| Model performance | 24 | Describe model performance | Model performance to be evaluated based on quality metrics described in methods.<br><br>Additionally, standard reporting of ML results should include confidence intervals, AUC and Balanced Accuracies (BAC), to allow for comparison of model performance with other published studies. | Pg8-11 |
| | 25 | How does the final model compare to other predictive tools? | Refer to model selection criteria as outlined in methods. | NA |
| | 26 | Bias and variance assessment | Prediction errors can be assessed with perturbation resampling, bootstrap resampling etc. | NA |
| | 27 | Model output interpretation | Where possible, report the variables used for prediction of outcome(s). | Pg12 |

| | | | | |
|---|---|---|---|---|
| | | | Report population subsets that were challenging, or easy, to predict. | |
| **DISCUSSION** | | | | |
| Clinical implementation | 28 | Describe how the model may be harnessed for improved patient care | Describe how the patient stands to benefit from; this may include savings in time, finances, side-effects.<br>Describe the most appropriate setting for application of the model. | Pg14-18 |
| Model limitations | 29 | Data format | State if there are any particular data format requirements, especially those that may hinder widespread use of the model in an appropriate setting. | Pg14-18 |
| | 30 | Justify choice of ML method | This may be related to purpose (eg interpretability), data characteristics (modality, quantity and quality), computational resources. | Pg14-18 |
| | 31 | Discuss potential bias in data sampling | | |
| | 32 | Discuss generalisability of the model | Does the model have applicability to other sample collection sites and particularly sites with different outcome distributions? Leave group out cross-validation can address site/group effects. | Pg14-18 |

#### SUPPLEMENTARY REFERERNCES

Akiba, T., Sano, S., Yanase, T., Ohta, T. and Koyama, M. 2019. Optuna: A Next-Generation Hyperparameter Optimization Framework. In: *The 25th ACM SIGKDD International Conference on Knowledge Discovery & Data Mining*. pp. 2623–2631.

Andreasen, N.C. 1984. *The Scale for the Assessment of Positive Symptoms*. Iowa City, IA: University of Iowa.

Andreasen, N.C. 1989. The Scale for the Assessment of Negative Symptoms (SANS): conceptual and theoretical foundations. *The British journal of psychiatry* 155(S7), pp. 49–52.

Quinn, T.P. et al. 2024. A primer on the use of machine learning to distil knowledge from data in biological psychiatry. *Molecular Psychiatry* 29(2), pp. 387–401. doi: 10.1038/s41380-023-02334-2.
